# Preventing premature deaths through polygenic risk scores

**DOI:** 10.1101/2024.12.26.24319670

**Authors:** Melisa Chuong, Deborah Thompson, Michael Weale, Fernando Riveros-McKay, Nilesh Samani, Daniel Wells, Vincent Plagnol, Gil McVean, Euan A Ashley, Peter Donnelly, Seamus Harrison, Jack W O’Sullivan

## Abstract

**Background:** Polygenic risk scores (PRS) have demonstrated predictive validity across a range of cohorts and diseases, but quantifying their clinical utility remains a challenge. As PRS can be derived from a single biological sample and remain stable throughout life, we explore the potential of PRS to optimize existing screening programs.

**Methods:** Via an integrated modelling approach, we quantified the potential clinical benefits arising from a knowledge of PRS across seven diseases with existing screening programs (abdominal aortic aneurysm, breast cancer, colorectal cancer, coronary artery disease, hypertension, prostate cancer, and type 2 diabetes). We identified individuals at high genetic risk (PRS OR>2) and very high genetic risk (PRS OR>3) and estimated the optimal screening ages for these genetically high risk individuals, based on the equivalent risk to population-level risk at recommended screening ages. We then leveraged published data on differential mortality and other outcomes, with and without screening-based interventions, to assess the potential benefits of tailoring screening age based on genetic risk. We also estimated the case enrichment ratio, which is a ratio of the percentage of cases in the high PRS risk group and in the total population.

**Findings:** Very high risk individuals reach the risk level associated with usual starting screening age on average 10.8 years earlier, high risk individuals 8.9 years earlier and reduced risk individuals (OR<0.5) 16.8 years later. During this time, case enrichment in the high risk group is between 1.7 and 3.0 depending on disease. Across all seven diseases, appropriate interventions following PRS-guided screening could reduce premature deaths in high risk individuals by 23.3%.

**Conclusion:** Knowledge of genetic risk, measured using PRS, has the potential to deliver substantial public health benefits when aggregated across conditions, and could reduce premature mortality by tailoring existing screening programs.

## Introduction

Common, chronic diseases account for over 90% of the $4.5 trillion annual healthcare spend in the US.^1^ There are a range of public health screening programs aimed at reducing the burden of these diseases, either via early detection (e.g. screening for asymptomatic cancers) or prevention (e.g. screening for cardiovascular disease risk). Most programs use simple risk factors for determining initial access to screening, notably age and sometimes sex. This approach leads to overtreatment in some,^2^ and undertreatment in others.^3^

Despite its widespread use, fixed age cutoffs fail to account for the considerable variability in individual risk, often missing high risk individuals who are younger than the minimum age threshold, whilst including lower risk individuals just because they are within the target age-group.^4^ Data on the effectiveness of age-based criteria in capturing true high risk populations are limited, and the current approach may inadvertently reduce the efficiency and equity of screening programs. Understanding the spectrum of risk across the population may allow better targeting of interventions to those most likely to benefit, and least likely to be harmed.

One promising avenue for improving risk stratification lies in genetic factors. Polygenic risk scores (PRS), which aggregate the effects of common genetic variants associated with a disease into a single score, have emerged as novel biomarkers that are stable throughout life and available for a broad spectrum of diseases.^5,6^ For many diseases, the strongest measurable risk factor is a PRS.^7^ Unlike other biomarkers, a single genetic test allows the construction of multiple PRSs that can be used to predict the risks of as many diseases as required.

In this study, we estimate the potential clinical benefits of tailoring current screening programs to genetic risk. Specifically, we calculate the proportion of the population who are at sufficient risk of disease to warrant earlier initiation, identify the age at which this earlier screening should occur, and quantify the mortality benefits of earlier, PRS-guided screening.

## Methods

### Screening diseases

We identified a list of seven common diseases for which age-related public health screening programs exist: abdominal aortic aneurysm, female breast cancer, colorectal cancer, coronary artery disease (which we define as a combination of either fatal coronary heart disease and/or myocardial infarction), hypertension, prostate cancer, and type 2 diabetes (see Supplementary Materials for more information). ‘Screening’ refers here either to screening for an existing condition to facilitate earlier therapeutic intervention, or to screening for risk to facilitate preventive intervention. For some diseases (abdominal aortic aneurysm, prostate cancer, type 2 diabetes), screening is only recommended in certain high risk groups, or following a patient-clinician discussion of benefits and harms based on the patient’s risk. We assume that possession of a high risk PRS value constitutes sufficient motivation for screening in this group (see Supplementary Materials).

### PRS and PRS effect sizes

We previously constructed and released the UK Biobank PRS Release v2 as a freely-available resource for UK Biobank researchers.^6^ The UK Biobank (UKB) is a UK based prospective cohort of ∼500,000 individuals, aged 40-69 when recruited between 2006 and 2010.^8,9^ We used the Standard PRS set, which was constructed from non-UKB training data only, as our source of high-performance PRS, along with their corresponding trait definitions. Briefly, the PRS were generated using meta-analyzed external genome-wide association study summary statistics. Per-individual PRS values were calculated as the genome-wide sum of the per-variant posterior effect size multiplied by allele dosage. All PRS were centered and standardized based on external reference data (1000 Genomes Project) to ensure means of approximately zero and standard deviations of approximately one within major genetically inferred ancestry groups. The same pipeline was used to construct a new PRS for abdominal aortic aneurysm. Disease phenotypes are defined using a combination of linked hospital episode statistics, cancer registry data and primary care data (see Supplementary Materials).

The UKB cohort was split into study training and test sets. Genetic ancestry was inferred using scores derived from principal components analysis of genetic data.^6^ The UKB training set was designed to include all participants with available PRS and non-European genetic ancestry (2867 with inferred East Asian (EAS) ancestry, 9543 with South Asian (SAS), 9486 with Sub-Saharan African (AFR), and 389 with Native/Indigenous American (AMR) ancestry), along with a random sample of 100,000 participants with European (EUR) ancestry. This UKB training set (122,285 individuals) was used to estimate the odds ratio per standard deviation (OR per SD) of each PRS in each ancestry group, using logistic regression.

Following removal of 31,300 UKB individuals who were related (up to third degree) with individuals in the training set, the remaining 332,664 individuals with available PRS were designated as an independent UKB testing set. The testing set consisted solely of individuals with inferred European ancestries, as the numbers with other ancestries were not large enough to allow useful training/testing splits. The UKB testing set was only used for two purposes: to calculate percentages of individuals at high genetic risk for multiple diseases; and to perform a sensitivity analysis to compare the primary results based on national incidence data with results based on observed case numbers.

Following previous practice,^7,10–12^ we defined individuals to be at ‘high risk’ of a disease if their PRS value was associated with an odds ratio (OR) >2 compared to the population mean risk. For ‘very high risk’, we used a PRS value associated with OR>3, and for ‘reduced risk’ we used a PRS value associated with OR<0.5 (Figure 1A). A detailed description of the calculation of risk groups is available in the Supplementary Materials.

**Figure 1.**
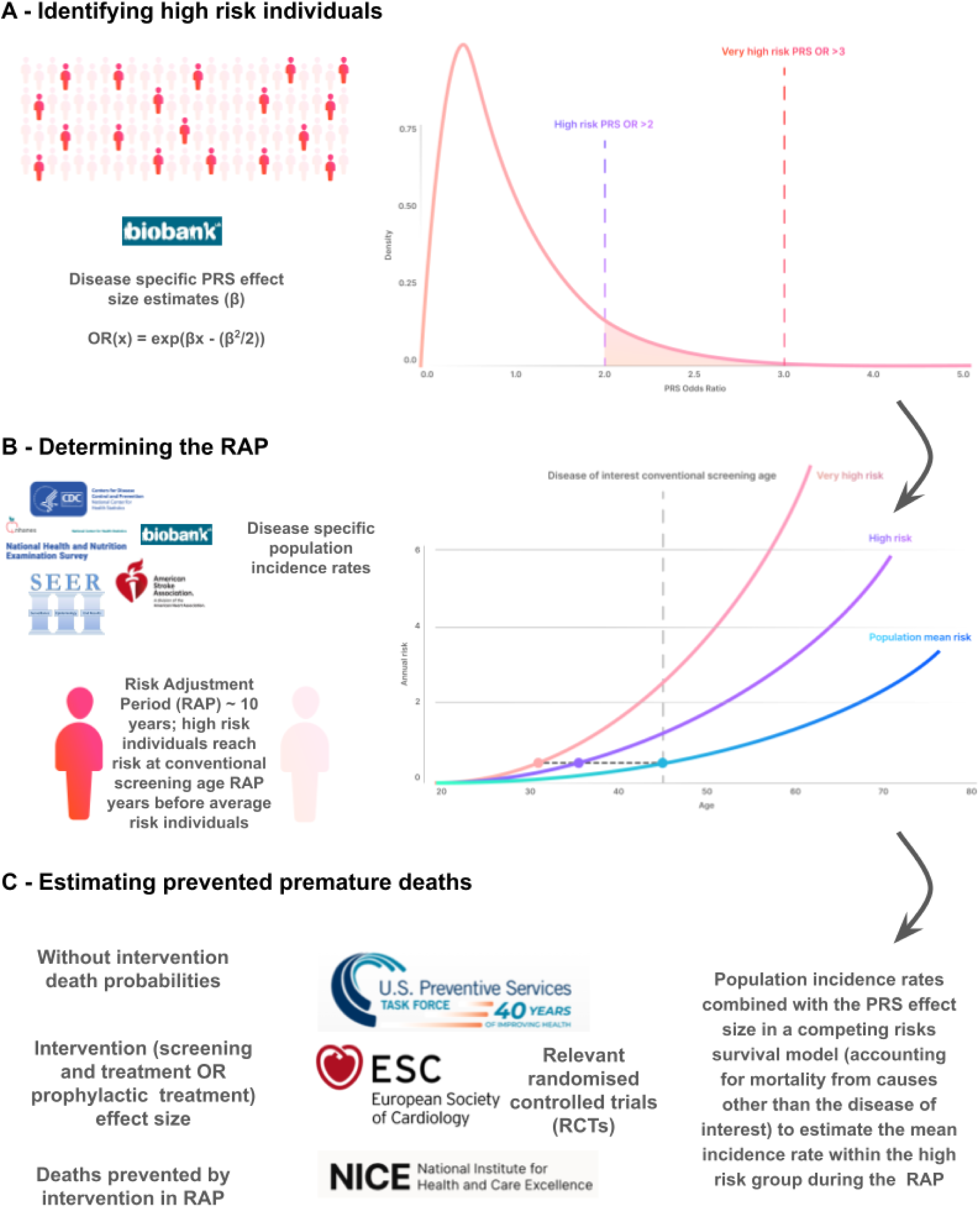
Overview of study methods. For each of seven screening diseases, we estimated the proportion of the population which could be considered to be at high genetic risk on the basis of their PRS (Panel A, see also “PRS and PRS effect sizes”). We then identified the age at which PRS-defined high risk individuals would reach the same disease incidence rate as that of the average population at the conventional starting age for screening, which defines the start of the risk advancement period (Panel B, see also “Risk advancement periods (RAP)”), and modelled the number of cases which would be diagnosed in these high-risk individuals between this earlier screening age and the current screening age (see “Estimation of events and deaths without PRS-guided screening”). From this, we estimated the number of cause-specific deaths which would result from cases diagnosed within this period, and hence the potential reduction in mortality which could be achieved by earlier intervention in this group (Panel C, see also “Quantification of clinical benefits of PRS-guided screening”).

### Risk advancement periods (RAP)

For each of the seven diseases, we extracted recommendations on screening and screening start age, predominantly relying on US based recommendations from the United States Preventive Services Task Force (USPSTF) (see Supplementary Materials). Following USPSTF recommendations, we only considered screening for abdominal aortic aneurysm in men.

For each disease, we then used published incidence data for the US population, stratified by age, sex and self-reported race/ethnicity, to calculate a set of baseline one-year incidence rates from age 0 to 80 years (see Supplementary Materials). Disease incidence data were obtained from a number of sources, and reflect the real world diagnosis rate at each age under current standard of care. We were unable to identify suitable US population incidence data for abdominal aortic aneurysm, so we instead used male incidence data derived from the UK Biobank. To estimate the incidence rate in each disease-specific high risk group, baseline incidence rates were multiplied by the mean relative risk within the high risk group compared to baseline, which was derived theoretically from the mean OR per SD in the relevant upper portion of the PRS distribution (using the equivalence between relative risk and OR that applies when baseline incidences are low, adjusting for bias in the transformation from PRS to OR (see Supplementary Materials), and assuming a constant relationship between PRS effect size and age).^13–15^

Following standard methodology for the construction of the risk advancement period,^16^ we then compared the incidence-by-age curves in order to find the age at which individuals in the high risk group reached the same level of risk as that found in the whole population at the conventional starting age for screening. We refer to this starting age as the PRS-guided screening age. The difference between the conventional screening age and the PRS-guided screening age defines the risk advancement period (RAP) (Figure 1B).^16^

Analyses were repeated for disease-specific very high risk groups (OR>3) and for reduced risk groups (OR<0.5). For reduced risk groups, the PRS-adjusted screening ages were later than the conventional screening ages, reflecting a negative RAP (i.e. a risk delay period).

For some diseases, the reduced risk group does not reach, at any age, the population risk at the conventional screening age. In this situation, calculations were based on taking the PRS-adjusted screening age to be 80 years, although we note that this does not reflect a realistic use-case.

The primary analyses used US incidence rates corresponding to White Non-Hispanic self-reported race/ethnicity, PRS effect size estimates corresponding to genetically inferred European ancestry, and PRS-adjusted screening ages that were calculated using baseline rates averaged over males and females, where applicable (sex-specific ages are reported in the Supplementary Materials). Sensitivity analyses explored the use of alternative conventional starting ages for screening, alternative US-based race/ethnicity for incidence rates and genetic ancestry for PRS effect size estimates, and alternative UK-based incidence rates derived either from external data sources or empirically from UK Biobank data.

### Estimation of events and deaths without PRS-guided screening

We used an integrated modelling approach to estimate events and deaths without PRS-guided screening. Population disease incidence data, stratified by age, sex and self-reported race/ethnicity and obtained from various national sources (see previous section and Supplementary Materials), were combined with estimates of mortality from other causes and with estimates of PRS effect size into a competing mortality survival model.^17^ The cumulative incidence function of this model was used to estimate the ‘without screening’ events, which is the number of diagnosed disease events, per 100,000 males and 100,000 females, that are predicted to occur without the imposition of a formal PRS-guided screening program, and without the additional interventions that would flow from such a program, during the RAP time interval between the earlier PRS-guided starting age for screening and the conventional starting age for screening. We note that the ‘without screening’ events are estimated from real world data, and therefore include any opportunistic screening, special screening and standard-of-care interventions that may occur during the RAP time interval. The cohort of 100,000 males and 100,000 females is modelled separately for each disease, and is assumed to be free of that disease at the start of the RAP period. We note that this is a conservative estimate of the true number of disease events for diseases where the purpose of screening is for early detection, because it is based on reported population incidence rates under current standard of care, and so does not include early-stage cases that might additionally be detected through earlier screening.

The without-screening events in the high PRS risk groups were found by rerunning the same competing mortality survival model, including a subdistribution hazard rate multiplier to represent the PRS effect size which is sampled from the appropriate top truncated portion of the PRS distribution. These without-screening disease events represent opportunities for early detection or preventive intervention (Figure 1C). As with the RAP calculation, we use the equivalence between relative risk and OR that applies when baseline incidences are low, we adjust for bias in the transformation from PRS to OR (see Supplementary Materials), and we assume a constant relationship between PRS effect size and age.^13,14^

The without-screening events in the high PRS risk groups were calculated separately for males and females, and the confidence intervals (CI) were derived from the 95% CI of the log(OR) per SD of the PRS, assuming that the PRS-guided screening age and the proportion of the population in the high risk groups are known without error, and ignoring uncertainty in the population baseline rates.

We estimated the ‘case enrichment ratio’ as the ratio of the expected case incidence in each high PRS risk group to the expected case incidence in the total population (equivalently, by rearrangement, it is the ratio of the percentage of high PRS risk group membership within cases to the percentage of high PRS risk group membership in the total population). For example, for breast cancer, 7.08% of all cases occurring during the RAP are in the high risk group, and 2.98% of the female population is in the high risk group, thus the case enrichment ratio is 2.37.

To estimate without-screening deaths, we conducted a literature search to identify large randomized controlled trials studying the efficacy of screening and screening-based interventions, and for which information on the number of disease-related events and deaths occurring in the control arms of the trial was available (see Supplementary Materials). From these data, we derived the conditional probability of disease-specific death, conditional on experiencing a disease event within the control arm. The without-screening events were multiplied by this conditional probability to give the estimated number of without-screening deaths per 100,000 males and 100,000 females, arising from events occurring within the RAP. Note that while these deaths are conditional on disease events that are diagnosed within the RAP, the deaths themselves can occur after the RAP time period (see next section).

### Quantification of clinical benefits of PRS-guided screening

We consider two kinds of clinical benefit: an ‘early detection’ benefit that arises from earlier screening, and thus, earlier therapeutic intervention for current disease, and an ‘early prevention’ benefit that arises from earlier preventive intervention that is administered to individuals at high risk due to their PRS. A literature search was conducted to identify randomized controlled trials on the effects of screening and screening-based interventions on relevant outcomes over sufficiently long follow-up time periods (see Supplementary Materials). From these data, we extracted relative risks describing the efficacy of screening and screening-based intervention, assuming the same level of compliance to screening and intervention as seen in the randomized controlled trials. For diseases where screening is for early detection, the relative risk is the relative mortality seen in the intervention versus control arms. For diseases where screening is for early prevention, the relative risk describes either the relative event incidence or relative mortality in the intervention versus control arms. Under both scenarios, the estimate of deaths expected under screening and screening-based intervention is obtained by multiplying the without-screening deaths by the relevant relative risk. The screening benefit arises either from improved survival outcomes following earlier detection (for diseases where screening is for early detection), or from reducing the disease event rate within the RAP time period (for diseases where screening is for early prevention).

When quantifying the combined benefit over all diseases, to avoid double-counting, we only included the benefit of ‘early detection’ and excluded the benefit of ‘early prevention’ for breast cancer. We did not estimate clinical benefits of delaying screening in the reduced risk groups as there are no randomized controlled trials or efficacy estimates for delayed screening.

We conducted a series of sensitivity analyses to explore the impact of altering various inputs on our modelling. These included: (1) using very high PRS risk (OR>3) to define the at-risk PRS groups; (2) using alternative conventional starting ages for screening; (3) using observed cases in the UK Biobank testing set during the RAP period, rather than modelling from separate sources of incidence and mortality data; and (4) modelling UK based incidence and mortality rates for a subset of diseases (see Supplementary Materials).

## Results

### Individuals at risk for multiple diseases

PRS effect size estimates (OR per SD) were determined in the UKB training set (Table S1) and used to define the disease-specific high risk and very high risk individuals in the UKB testing set. Of the 332,664 individuals in the UKB testing set, there were 83,950 individuals at high risk (PRS OR>2) and 18,956 individuals at very high risk (PRS OR>3) for at least one of the seven diseases for which current screening programs exist (Table 1), corresponding to approximately 25% and 6% of the cohort, respectively.

**Table 1.** Count (and percentage) of individuals in the UKB testing set at high (PRS OR>2) and very high risk (PRS OR>3) for the seven screening diseases.

| Number of Diseases | At High Risk | At Very High Risk |
| --- | --- | --- |
| 0 | 248714 (74.8%) | 313703 (94.3%) |
| 1 | 73119 (22.0%) | 18520 (5.6%) |
| 2 | 9905 (3.0%) | 436 (0.1%) |
| 3 | 868 (0.3%) | 5 (0%) |
| 4 | 55 (0.0%) | 0 (0%) |
| 5 | 3 (0.0%) | 0 (0%) |
| At least one | 83950 (25.2%) | 18961 (5.7%) |

### High PRS individuals are predicted to reach equivalent risk earlier - Risk Advancement Period (RAP)

For high PRS risk groups (PRS OR>2), we predict that high risk individuals would reach the target incidence rate between 4 and 22 years (mean 8.9 years) before the conventional starting age for screening, depending on disease (Figure 2, Table S2). The disease with the largest RAP for high risk individuals was hypertension (Figure 2, Table S2). A similar pattern is seen for the very high risk groups (OR>3), with RAP ranging from 5 to 22 years (Table S2) (mean 10.8 years). Individuals with a reduced PRS risk (OR <0.5) are predicted to reach population level risk 6 to 40 years later (mean 16.8 years) than the conventional starting age for screening; for prostate cancer (based on a screening start age 55) and hypertension, we predict that the reduced risk groups will never reach the target incidence (Figure 2, Table S2).

**Figure 2.**
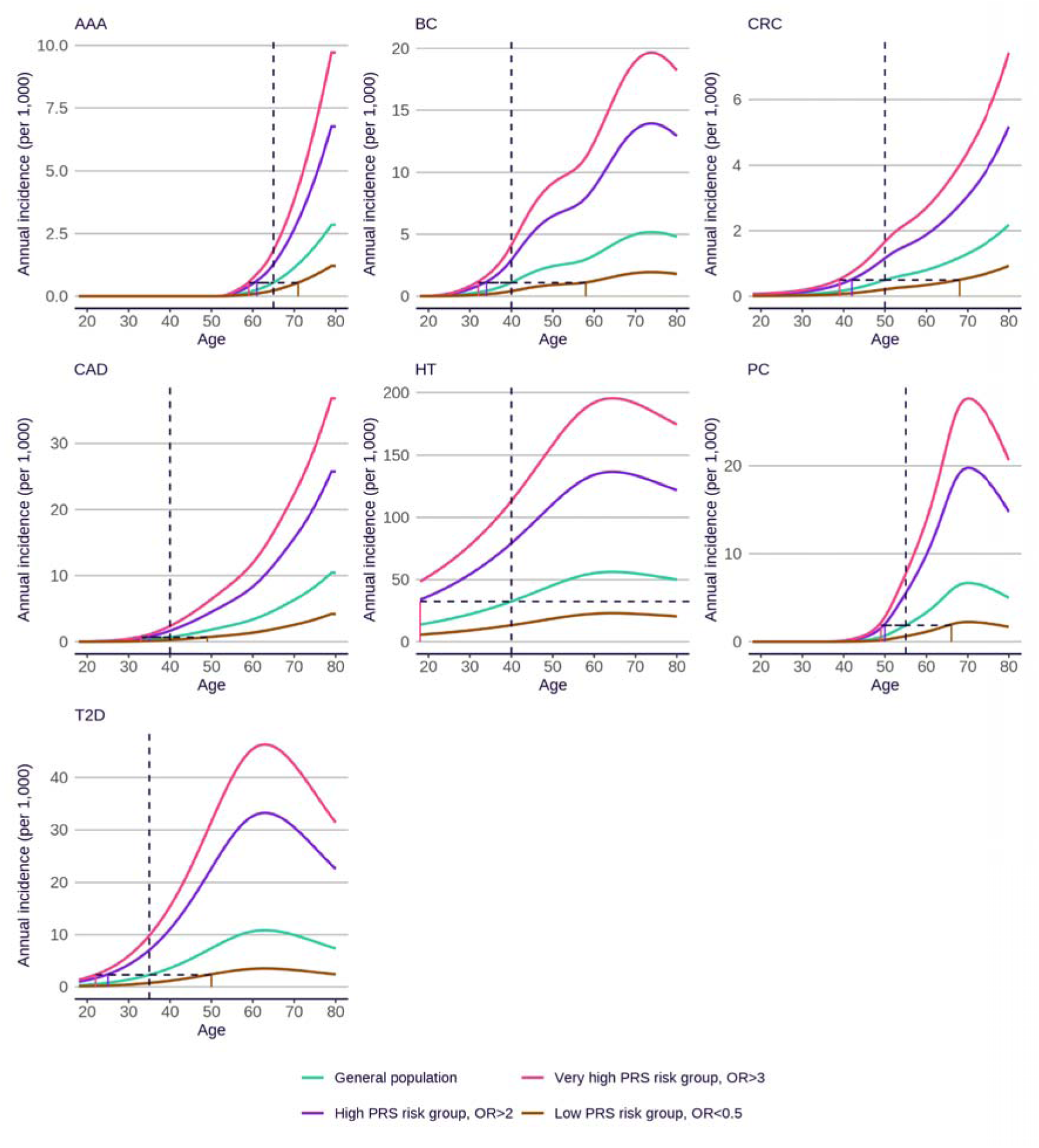
Annual incidence with age for individuals at high (PRS OR>2), very high (PRS OR>3), and reduced risk (PRS OR<0.5), compared to the population mean. Incidence data are based on US White Non-Hispanic self-reported race/ethnicity, and PRS effect sizes are based on European genetic ancestry (see Supplementary Materials for figures based on alternative sources). Incidences are sex-averaged, apart from prostate cancer (male), breast cancer (female) and abdominal aortic aneurysm (male). Vertical dotted lines indicate the conventional screening start age. Horizontal dotted lines indicate the RAPs for different PRS-based risk groups. AAA = abdominal aortic aneurysm; BC = female breast cancer; CRC = colorectal cancer; CAD = coronary artery disease; HT = hypertension; PC = prostate cancer; T2D = type 2 diabetes.

Alternative conventional starting ages for breast cancer, colorectal cancer and coronary artery disease were explored for sensitivity analyses, with results summarized in Figure S1 and Table S2. The analyses were also repeated, replacing the population incidence rates with those for Black Non-Hispanic self-reported race/ethnicity and using a PRS effect size estimated in the genetically-inferred African-ancestry UKB training set (Figure S2, Figure S3, Table S1). Here the PRS effect sizes are smaller, meaning that fewer individuals have a PRS OR high enough to be in a high risk group. However, the slightly lower average OR within each high risk group leads to only a relatively small reduction in the length of the RAP for these diseases. For some diseases (breast cancer, colorectal cancer, prostate cancer and type 2 diabetes), we also repeated RAP calculations based on UK-based conventional starting ages, UK self-reported White race/ethnicity incidence rates and European ancestry PRS effect sizes, which yield similar RAP durations (Figure S4).

### The clinical benefits of PRS-guided screening and treatment

For the seven diseases with existing screening programs, we determined the number of disease-specific deaths that could be prevented by offering earlier PRS-guided screening or intervention to individuals who are at risk due to their PRS. For each disease, we identified the probability of disease associated death once diagnosed as a case within control groups of relevant randomized controlled trials; the appropriate screening or screening-based intervention based on guidelines; and the corresponding screening or intervention efficacy (relative reduction in relevant outcome) based on randomized controlled trials (Table 2). For breast cancer, we identified efficacies for both screening (mammography) and for risk-lowering preventive intervention (tamoxifen).

**Table 2.**
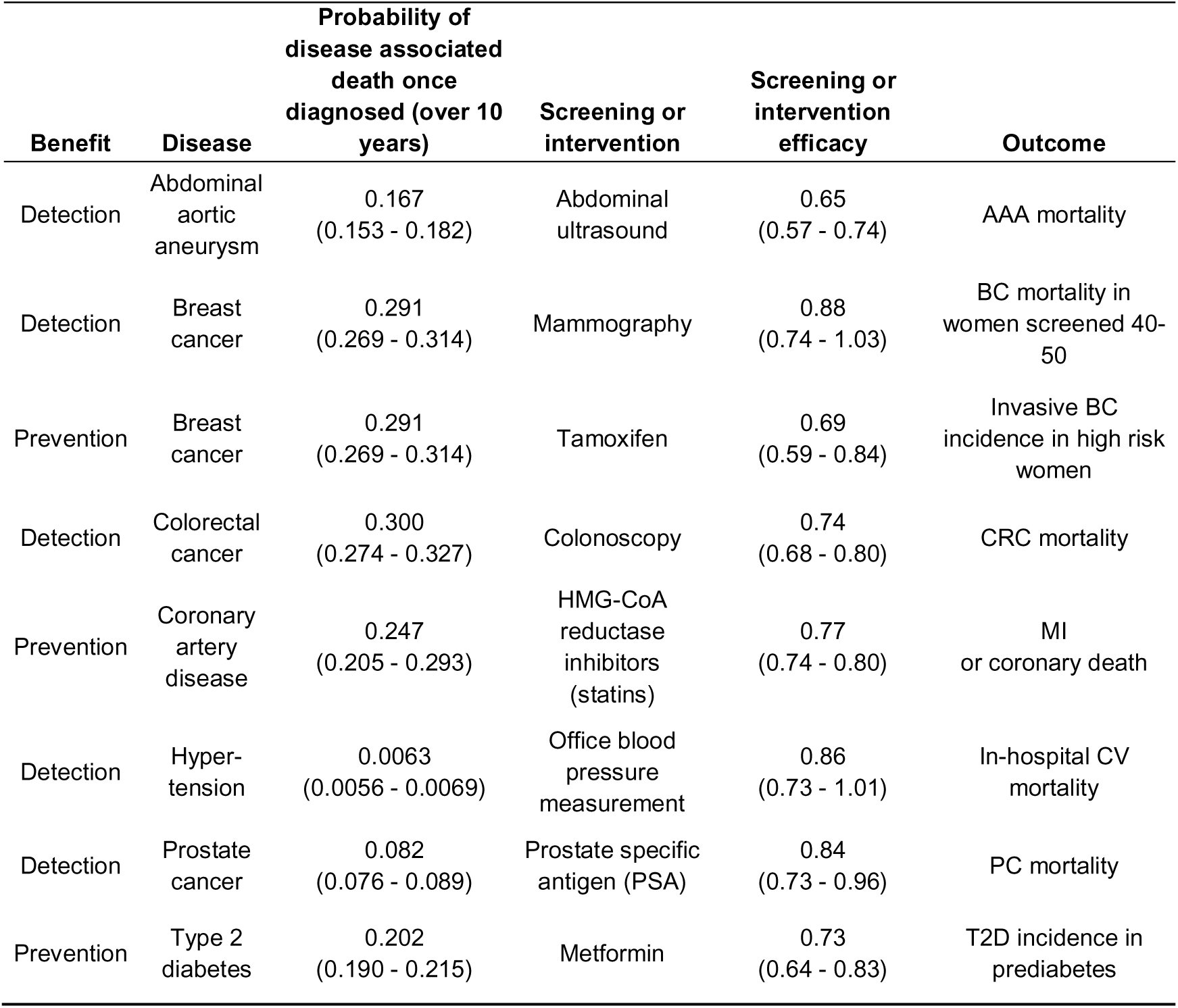
Probability of disease associated death (with 95% CI), guideline-based screening or intervention, the screening or intervention efficacy in terms of relative outcome reduction (with 95% CI), and the relevant outcome affected, for the seven screening diseases under early detection or early prevention benefit. MI = myocardial infarction; for other disease abbreviations see Figure 2.

The early detection and early prevention benefits are displayed respectively in Tables 3 and 4, using a PRS threshold of OR>2 to define disease-specific high risk groups. Across the seven diseases, between 2.9% (colorectal cancer) and 10.0% (type 2 diabetes) of the population are identified as being at high risk due to their PRS. Within these high risk groups, there is a case enrichment within the RAP of between 1.7 (hypertension in men) and 3.0 (type 2 diabetes in men) compared to the general population, resulting from their increased genetic risk (Figure 3). If relevant screening and screening-based interventions were applied within these high risk groups, we estimate that between 12% (breast cancer) and 35% (abdominal aortic aneurysm) of disease-specific deaths within these high risk groups could be prevented via early detection (Table 3), and between 23% (coronary artery disease) and 31% (breast cancer) could be prevented via early preventive intervention (Table 4). Summing across both types of benefit (but counting only the early detection benefit for breast cancer to avoid double counting), we estimate that, in a population of 100,000 individuals of each sex, 6387 disease cases could potentially occur within the RAP in individuals with a high PRS risk for the relevant disease. These cases, which fall outside of current screening ages, could be expected to lead to 256 deaths. If a PRS-guided early detection and/or early prevention strategy was adopted, 60 of these deaths (23.3% of disease-specific deaths in the respective high risk groups) could be saved.

**Figure 3.**
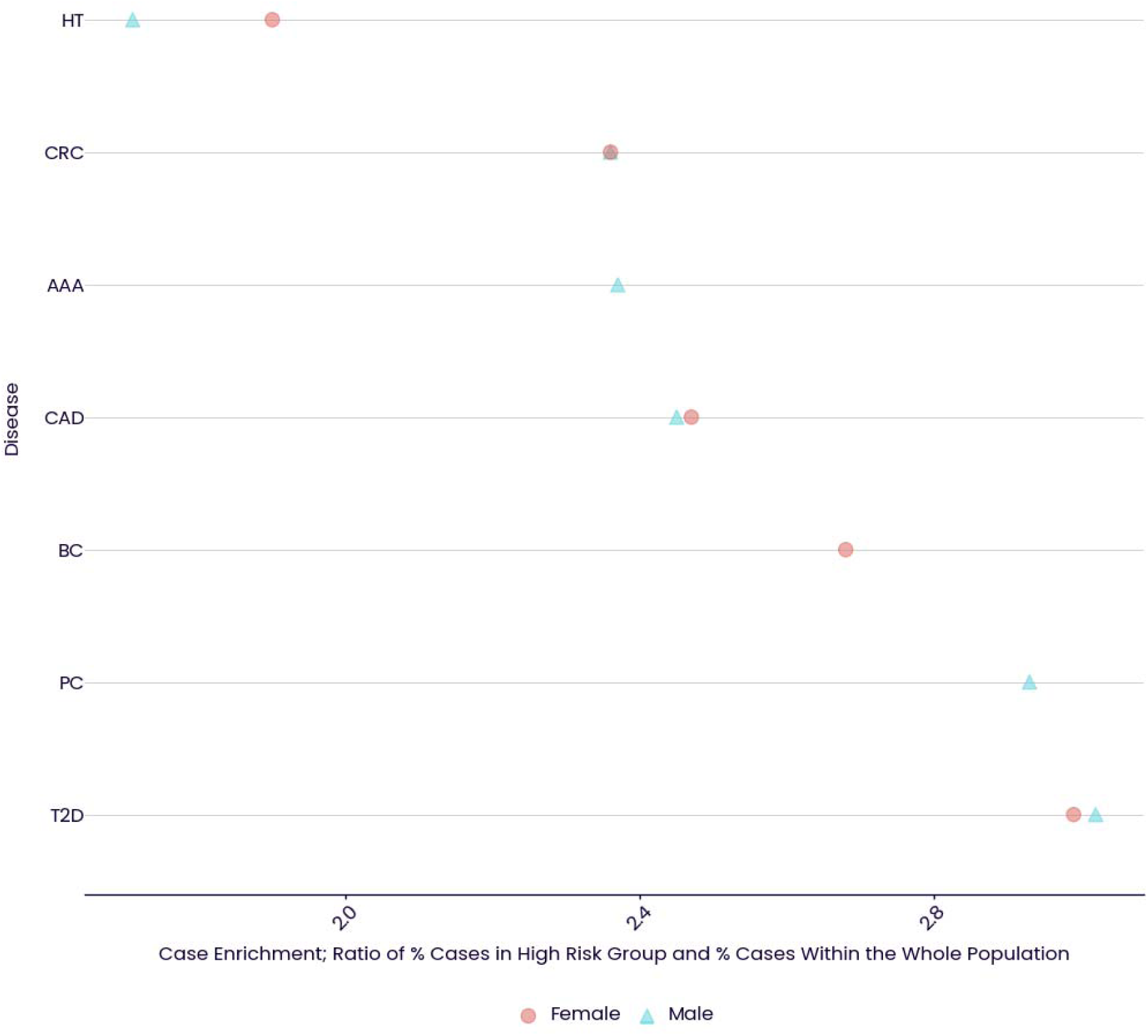
Case enrichment (ratio of the percentage of cases in high risk (PRS OR>2) individuals compared to that in the whole population), in men and women for the seven screening diseases.

**Table 3.**
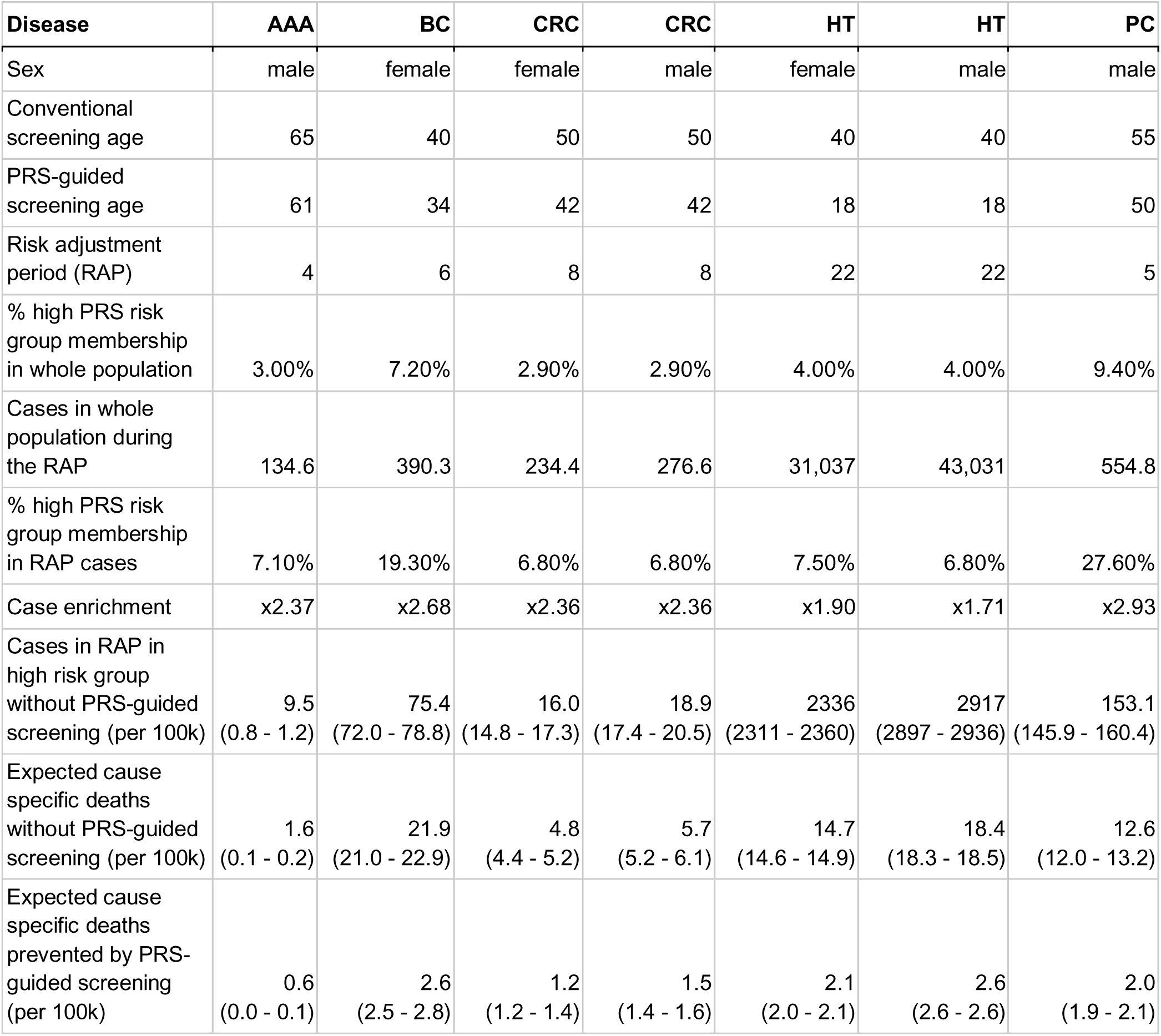
Estimates of clinical benefits of PRS-guided screening leading to early detection benefit. Numbers in the bottom three rows report expected numbers of cases within the RAP in the high PRS risk group, expected deaths from those cases without screening, and expected deaths prevented by PRS-guided screening, per 100k population of the corresponding sex. Numbers in brackets are 95% confidence intervals. For disease abbreviations see Figure 2.

**Table 4.** Estimates of clinical benefits of PRS-guided screening leading to early prevention benefit. Numbers in the bottom three rows report expected numbers of cases within the RAP in the high PRS risk group, expected deaths from those cases without screening, and expected deaths prevented by PRS-guided screening, per 100k population of the corresponding sex. Numbers in brackets are 95% confidence intervals. For disease abbreviations see Figure 2.

| **Disease** | **BC** | **CAD** | **CAD** | **T2D** | **T2D** |
| --- | --- | --- | --- | --- | --- |
| Sex | female | female | male | female | male |
| Conventional screening age | 40 | 40 | 40 | 35 | 35 |
| PRS-guided screening age | 34 | 35 | 35 | 25 | 25 |
| Risk adjustment period (RAP) | 6 | 5 | 5 | 10 | 10 |
| % high PRS risk group membership in whole population | 7.20% | 4.40% | 4.40% | 10.00% | 10.00% |
| Cases in whole population during the RAP | 390.3 | 117.3 | 324.9 | 1513.1 | 1192.3 |
| % high PRS risk group membership in RAP cases | 19.30% | 10.90% | 10.90% | 30.00% | 30.20% |
| Case enrichment | x2.68 | x2.47 | x2.45 | x2.99 | x3.02 |
| Cases in RAP in high risk group without PRS-guided screening (per 100k) | 75.4<br>(72.0 - 78.8) | 12.8<br>(12.2 - 13.4) | 35.3<br>(33.7 - 37.0) | 453.2<br>(440.2 - 466.3) | 360.0<br>(349.6 - 370.6) |
| Expected cause specific deaths without PRS-guided screening (per 100k) | 21.9<br>(21.0 - 22.9) | 3.2<br>(3.0 - 3.3) | 8.7<br>(8.3 - 9.1) | 91.5<br>(88.9 - 94.2) | 72.7<br>(70.6 - 74.9) |
| Expected cause specific deaths prevented by PRS-guided screening (per 100k) | 6.8<br>(6.5 - 7.1) | 0.7<br>(0.7 - 0.8) | 2.0<br>(1.9 - 2.1) | 24.7<br>(24.0 - 25.4) | 19.6<br>(19.1 - 20.2) |

For example, in a cohort of 100,000 women who were without breast cancer at the start of the RAP, the 7,200women in the high PRS risk group for breast cancer (7.2% of the cohort) could begin screening for early detection at age 34, instead of at the conventional age of 40 (Table 3). Among these 7,200 high-risk 34 year olds, we could expect to see 75 breast cancer diagnoses between age 34 and 40. Assuming that 29% of cases would lead to breast-cancer related death (Table 2),^18^ these cases could result in 22 deaths. The UK Age randomized control trial reported a 12% reduction in breast cancer mortality associated with mammographic screening between ages 40-49 (as opposed to the UK standard screening age of 50 years).^18^ We hence estimate that 2.6 deaths could be prevented by PRS-guided screening in a cohort of this size.

We performed various sensitivity analyses to explore the impact of applying alternative modelling scenarios. Altering conventional starting age for screening tended to increase PRS-guided screening benefit if the age was later, and decrease it if the age was earlier (Table S3). Restricting interventions to individuals with a very high risk PRS (OR>3) resulted in longer risk advancement periods, but a more modest overall impact, because the earlier interventions would be offered to fewer people (e.g. 1.6% of women at very high risk of breast cancer, as opposed to 7.2% at high risk). Over all seven diseases, we estimate that 28 disease-specific deaths in the very high risk groups could be prevented in a population of 100,000 individuals of each sex, combining early detection and early prevention benefits (Tables S4 and S5). Using UK-based disease incidences, either using empirical case data from the UK Biobank testing set (Table S6 and S7) or using national data sources (Table S8), generated only modest differences in results.

## Discussion

Healthcare systems continue to invest large amounts of resources into screening for many common diseases. In order to optimize cost-effectiveness and minimize harms from overtreatment, access to screening for early detection or prevention is targeted at population groups for whom the benefits are deemed to be greatest. This targeting is achieved using a very minimal number of predictors, typically age and sex. As these are relatively crude predictors of risk, a reliance upon these factors can lead to overtreatment in some groups, and undertreatment in others. Here we consider using an additional immutable risk factor, PRS, to support stratification of screening and screening-based intervention opportunities. Specifically, we focus upon the potential clinical benefit of earlier intervention based upon genetic stratification across a range of important and common diseases.

Our paper shows three main results: 1) a single biological sample, obtained once, can predict the risk of many important common diseases, and can identify a substantial proportion of the population at high risk; 2) PRS can be used to propose rational starting ages for early screening in high risk people; and 3) if PRS-guided interventions were implemented, premature mortality could be substantially reduced.

First, we show that PRS can predict the risk of seven common diseases (abdominal aortic aneurysm, female breast cancer, colorectal cancer, coronary artery disease, hypertension, prostate cancer, and type 2 diabetes), and that a quarter of the population is at high risk of at least one of these diseases. PRS has the advantageous property that data from a single test can be obtained once at an early age, in principle from birth, and can capture risk simultaneously across many diseases.

Second, regarding screening for these seven diseases, we show how PRS information could be used to propose rational earlier screening start times for people at high risk. Compared to a strategy offering earlier invitations to individuals with a high PRS, we show that current screening misses opportunities to intervene in 6387 disease-specific cases per 100,000 individuals of each sex. Under current practice, notwithstanding infrequent familial cases detected by cascade screening,^19,20^ most of these cases would either be detected at later symptomatic stages, where therapeutic interventions typically have poorer outcomes, or would manifest without the presence of preventive interventions that could have reduced their risk of occurring.

Compared to when conventional screening begins, we predict that high risk individuals (PRS OR>2) would reach the equivalent population disease incidence rate around a decade earlier (ranging from 4 to 22 years across diseases). Hypertension (using a conventional starting age of 40) and breast cancer (using a conventional starting age of 50) were the diseases with the longest risk advancement periods. Conversely, reduced risk individuals (OR < 0.5) reached the equivalent population disease incidence rate on average more than a decade after conventional screening begins. Together, this demonstrates that using age alone is a relatively crude way to stratify population health programs.

Third, we show that premature, disease specific deaths that occur before conventional screening occurs are potentially preventable with PRS-guided interventions. Across our included diseases, our results suggest that 256 disease-specific deaths (per 100,000 individuals of each sex) would occur in the corresponding high risk groups before conventional screening occurs, and 23% of the disease-specific deaths within these high risk groups could be prevented with PRS-guided interventions. We estimate that the highest proportions of preventable deaths during the risk advancement period are for abdominal aortic aneurysm, breast cancer, and type 2 diabetes, with respectively an estimated 35%, 31% and 27% of disease-specific deaths preventable with PRS-guided interventions. In absolute terms, we found that type 2 diabetes, hypertension, and breast cancer had the potential for the largest number of deaths saved by this approach.

Our estimate of the absolute number of deaths prevented is small (60 per 100,000 individuals of each sex), but identifying and preventing mortality in younger adults is likely to lead to large improvements in quality adjusted life years. More broadly, earlier intervention would be associated with other benefits, beyond mortality reduction, including less aggressive (and less expensive) treatments, likely resulting in fewer side-effects. Identifying high risk individuals or those with diseases earlier also has the potential to reduce events such as non-fatal myocardial infarction and stroke. Indeed, in absolute terms, the reduction in non-fatal events is likely to be greater.

An alternative to PRS-informed intervention programs would be to lower the screening-invitation age for the whole population. However, we note this would involve considerably more resources. For example, the RAP period for breast cancer (using a conventional starting age of 40) is 6 years, with 7.2% of females in the high PRS risk group. Biennial screening for this high risk group (i.e. 3 extra screens) would be equivalent to 0.21 extra screens for the whole female population. Adding even just one additional screen for the whole population would result in nearly 5 times as many total screens as targeting 3 extra screens to the high PRS risk group.

The existing literature regarding PRS largely examines prediction of lifetime disease susceptibility,^10,21^ and integration of PRS into existing clinical risk scores.^10,22,23^ However, there are only a few studies concerning the identification of earlier conventional screening ages with PRS^21,24,25^, or predicting cumulative disease burden across a number of diseases using PRS.^26^ Our 8-year advancement period for colorectal cancer is broadly in line with the 5- and 11-year risk advancement periods estimated for the 80-99% and >99% PRS groups modelled in the Finnish population.^25^ Our results are also congruent for men compared to the study of Chen et al,^24^ who estimated personalized screening ages for men and women in UK Biobank in the highest PRS decile for colorectal cancer, but not for women (we predicted women would reach the same disease incidence rate at 34 whereas they predicted 48).

Reasons for this difference may include the use of different conventional screening ages (50 versus 55 years), and also our use of national sources of incidence data to support our estimates. Jermy et al examined the ability of PRS to estimate PRS-guided screening ages for breast cancer and type 2 diabetes.^21^ This paper used a different methodology, comparing PRS percentiles, and used the old USPSTF breast cancer screening age (50), and found that women at the top 5% PRS risk would be eligible for breast cancer screening at 43 years. In contrast, we used the new USPSTF breast cancer screening age of 40 and found a PRS-guided screening age of 34 years. For type 2 diabetes, Jermy et al found people at the top 5% PRS risk for type 2 diabetes would be eligible for screening at 29 years, whereas our results suggest 25 years of age. However, Jermy et al did not use USPSTF guidelines to define conventional screening ages, and did not use national US incidence data to support estimates. Widen et al examined the ability of PRS to identify people at high risk across a number of diseases.^26^ These authors constructed polygenic health indexes, and estimated the ability of PRS to predict lifetime disease burden when combining PRS across a number of diseases. Our study quantifies the number of people at high and very high risk for a number of diseases and then estimates associated mortality estimates.

### Assumptions and limitations

The findings of this study are based on an integrated modelling approach, combining model parameters from a wide variety of different sources. This approach introduces a number of assumptions at various stages of the modelling process, which we consider here.

The estimates of PRS effect size (the odds ratio per standard deviation of PRS) were obtained from UK Biobank. This assumes a robustness of PRS effect size to known health biases in UK Biobank, to differences in disease prevalence between the UK and US, and to potential differences in the way diseases are recorded or defined between the UK Biobank and the target US setting. We used ICD10 codes wherever possible to rely on international coding standards, but accept that application of these codes may vary between the UK and US. Some diseases, such as type 2 diabetes, are not well captured by ICD10 codes in a hospital setting, because they are typically diagnosed and treated within a primary healthcare setting. For these diseases, we used primary healthcare data from UK Biobank to define the disease (see Supplementary Materials). Studies have noted generally consistent PRS effect size between target populations of the same ancestry,^27,28^ and that health biases in UK Biobank do not necessarily impact the valid assessment of exposure-disease relationships.^29^

We also assumed that PRS effect sizes are constant with age. Studies have noted a small but consistent tendency towards larger PRS effect sizes in younger age groups.^6,30^ To the extent that this effect holds, our modelling will underestimate the effect of PRS-guided screening on deaths prevented in younger age groups. We also assumed that a PRS effect size calculated as an odds ratio can be applied as a hazard ratio in subsequent modelling. We have shown elsewhere that, in the context of UK Biobank, these two ways of measuring effect size are largely equivalent.^6^

US and UK national disease incidence and mortality data were obtained from a variety of recognised sources (see Supplementary Materials), but nevertheless these can be subject to potential uncertainties associated with their sampling and recording methods.

Estimates of the conditional probability of disease-specific death under no screening, given that a person is a diagnosed case for that disease, were obtained from the control arms of relevant randomised controlled trials. Estimates of screening or intervention efficacy were typically obtained from the same randomised controlled trials. These trials were conducted in a variety of settings, both in and outside the USA, with a range of trial conditions and follow-up times. Our modelling assumes that events and deaths in the control arms follow real world practice and standard of care during the RAP periods; that events and deaths in the screening or intervention arms accurately reflect real world implementation of that screening or intervention; that follow-up times are sufficiently long to capture pertinent disease-specific death events (we note that our modelling assumes that diagnosed case events must occur with the RAP periods, but disease-specific deaths caused by those case events can occur afterwards); and that both the conditional probability of death and efficacy benefit of screening is the same in people with high PRS risk as it is in the general population. We note this last point may sometimes be a conservative assumption - for example, there is evidence of an increased efficacy of statins in people with high PRS risk.^31,32^ The control arms of these randomised controlled trials are typically conducted on older individuals: we assume that both the conditional probability of death and the screening and intervention efficacies and modalities are the same for events diagnosed in younger people in the RAP periods. We note that optimal screening modalities in younger people may differ - for example, ultrasound may be more efficient for screening for breast cancer in younger women, which carries its own costs and characteristics.

We performed our modelling separately for each disease, and then summed the impact on deaths as a last step. This approach requires a low per disease death rate (to avoid issues relating to double counting), a requirement which was met by the relatively young age group in which our modelling was performed, and assumes independence between diseases. The confidence intervals we provide on deaths prevented per disease only take uncertainty on PRS effect size into account, and so do not reflect all sources of uncertainty.

Our primary analyses present results for individuals of European ancestry and White race/ethnicity, using guidelines and modelling tailored to the USA. However, we present various sensitivity analyses to indicate that PRS-guided screening could also be beneficial to individuals of non-European ancestry and race/ethnicity, in the UK as well as the US, and using alternative starting ages for conventional screening and alternative thresholds for defining the at-risk PRS group. Our analyses consider one of the simplest possible alternatives to a single conventional starting age for screening, which is to propose a single alternative starting age for individuals declared to be at risk due to their PRS. We note that a PRS affords a quantitative estimate of a person’s disease risk, and that therefore alternative schemes could be devised in which the screening starting age is individually tailored to a person’s PRS score, perhaps combined with other risk factors such as their sex (although we note there could be administrative complexities associated with such schemes). PRS-guided screening could also be applied to the consideration of delayed starting ages for individuals with low PRS, or to the consideration of tailored screening stopping ages. These alternative schemes are fruitful areas for future research. We also note that our simple PRS-only approach assumes that other risk factors, such as rare pathogenic variants for breast cancer, or such as the use of clinical risk tools for screening for high risk in coronary artery disease and type 2 diabetes, are dealt with via other pathways. We did not carry a formal cost-benefits analysis of our PRS-guided scheme, but note this would be a priority for future work.

In summary, our modelling approach applies a number of assumptions and simplifications, which influence the accuracy of our calculations. We carried out various sensitivity analyses, all of which demonstrated a benefit to PRS-guided screening, but inevitably these analyses could not cover all possible eventualities.

### Conclusions

Collectively, our results show the power of PRS across multi-disease risk prediction and precision-guided screening, with the potential to save premature, preventable deaths in younger adults. When considered in combination with prior work showing the feasibility of integrating PRS into primary care,^33^ our results show a promising clinical use case for PRS. Our results have implications for patients, clinicians, researchers and policy makers. For clinicians, PRS are increasingly available both via direct to consumer tests and within healthcare systems themselves.^34^ We anticipate our data may provide some guidance, by providing some data to support conversations regarding risk and screening for patients presenting to clinics with PRS results. For patients, our results show that PRS should be considered a risk factor that may help guide health behaviors. For policy makers, our results highlight the use of PRS as a risk factor that can improve screening. PRS can enhance accuracy and potentially identify people who are being missed by conventional screening programs.

## Data Availability

All UK Biobank data is publicly available upon application. We have made the polygenic risk scores used in this study publicly available.

https://github.com/genomicsplc/clinical_benefits

---

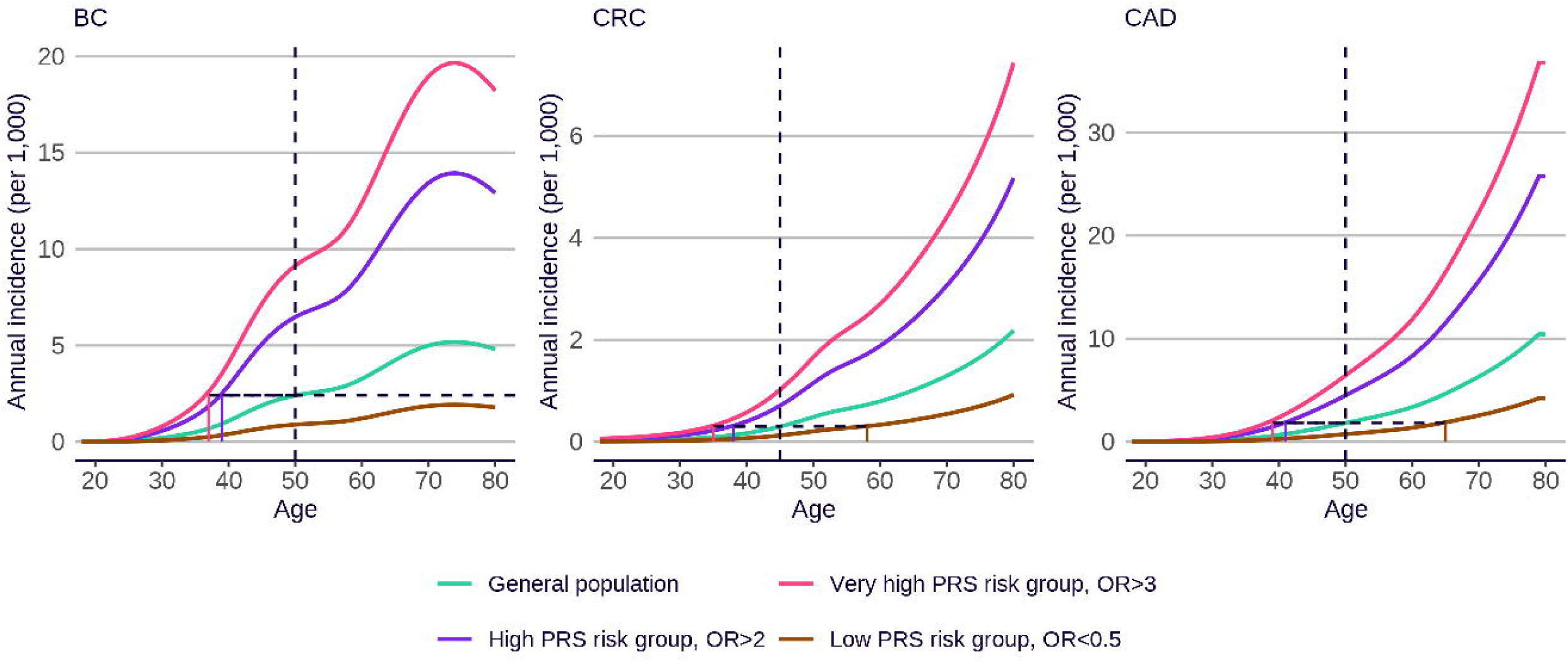

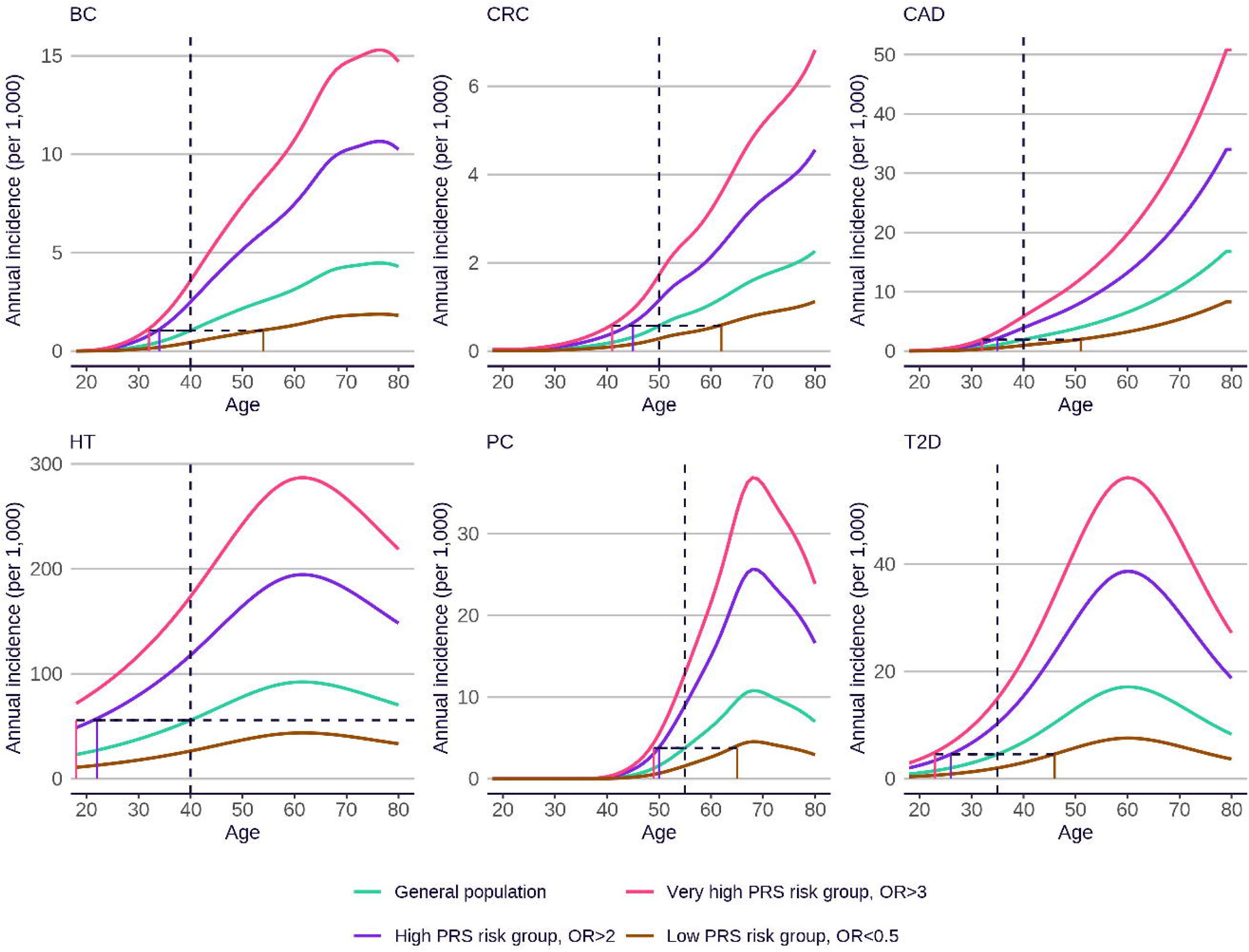

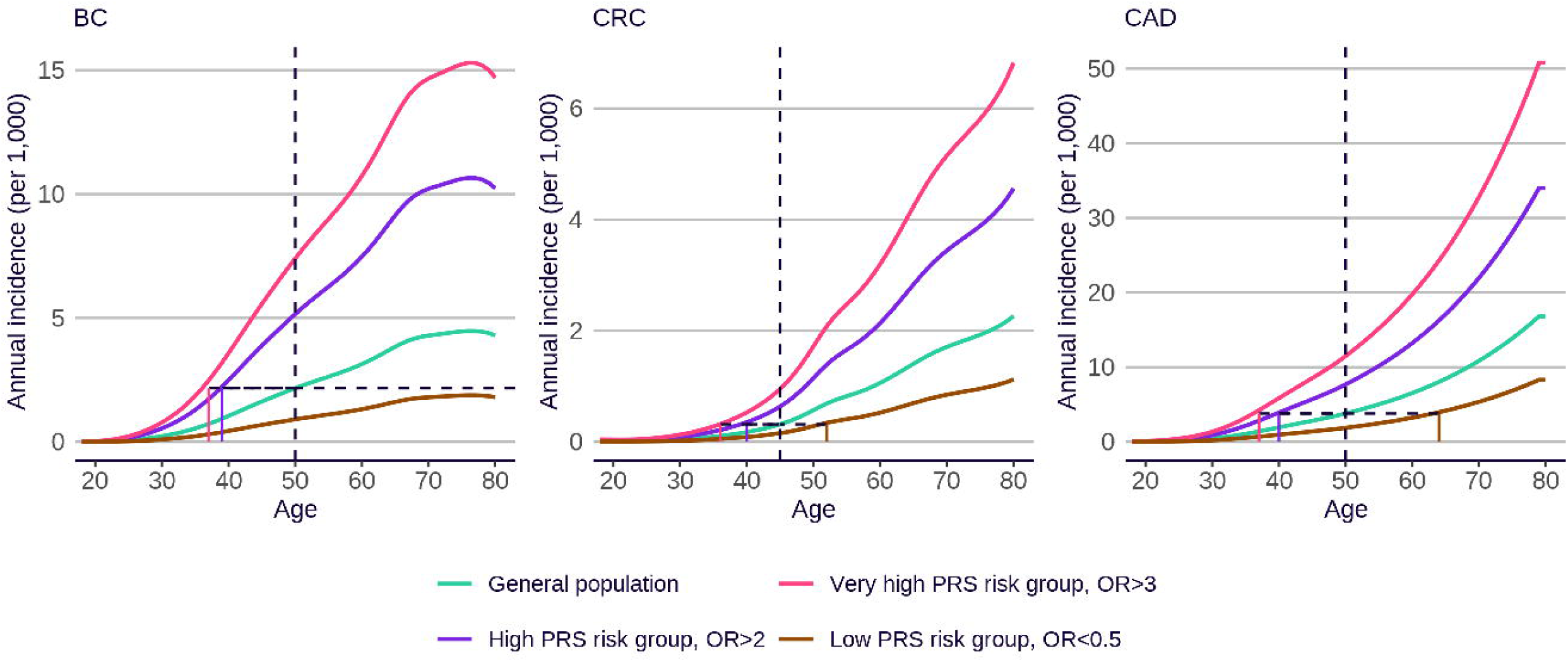

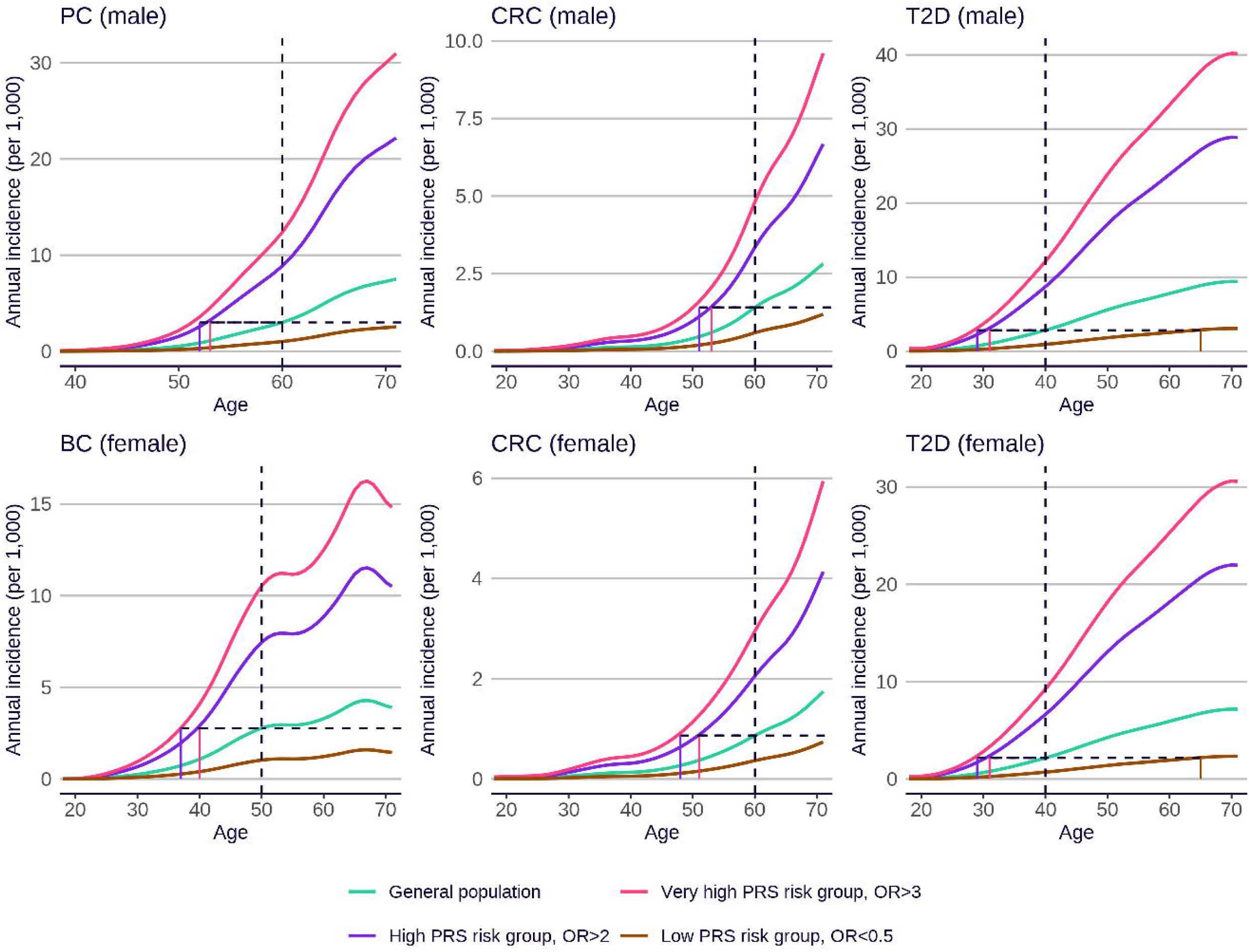

