## Supplementary material for "Preventing premature deaths through polygenic risk scores": Chuong_Preventing_premature_deaths_through_polygenic_risk_scores_Jul2025_SI.pdf

### Supplementary Materials

#### Supplementary Methods

##### Selection and review of screening diseases and guidelines

To identify relevant diseases subject to population-based screening, we performed a multi-faceted approach combining a systematic search and expert clinical knowledge. For the systematic search, we searched pubmed with MeSH terms ‘Mass Screening’, AND ‘Humans’ AND ‘Adults’ combined with domain-specific terms: [‘Early Detection of Cancer’ AND ‘Cancer], [‘Cardiovascular Diseases* / prevention & control’]. We also searched relevant global clinical screening bodies identified by clinical experts (JOS, SH). We focussed on screening guidelines intended for use in the USA by the United States Preventive Services Task Force (USPSTF). We provide guidelines from other relevant bodies where relevant. We excluded guidelines relevant only to individuals carrying rare, high-risk mutations (e.g. *BRCA1/2* or Lynch syndrome genes). Summaries of these guidelines are provided below.

**Abdominal aortic aneurysm**

USPSTF guidelines [1] state that: “The USPSTF recommends 1-time screening for abdominal aortic aneurysm (AAA) with ultrasonography in men aged 65 to 75 years who have ever smoked.”

Smoking is a risk factor for AAA. We assume that selective screening for AAA would also be motivated for men at high risk of AAA due to their PRS. It has been shown that risk inferred from a high PRS approaches that of the risk from smoking [2,3].

**Female breast cancer**

USPSTF guidelines [4,5] state that: “The USPSTF recommends biennial screening mammography for women aged 40 to 74 years” and “The USPSTF recommends that clinicians offer to prescribe risk-reducing medications, such as tamoxifen, raloxifene, or aromatase inhibitors, to women who are at increased risk for breast cancer and at low risk for adverse medication effects.”

**Colorectal cancer**

USPSTF guidelines [6] state that: “The USPSTF recommends screening for colorectal cancer in all adults aged 50 to 75 years.”

**Coronary artery disease**

USPSTF guidelines [7] state that: “The USPSTF recommends that clinicians prescribe a statin for the primary prevention of CVD for adults aged 40 to 75 years who have 1 or more CVD risk factors (i.e. dyslipidemia, diabetes, hypertension, or smoking) and an estimated 10-year risk of a cardiovascular event of 10% or greater.”

The American Heart Association (AHA) and American College of Cardiology (ACC) guidelines [8] state: “Adults who are 40 to 75 years of age and are being evaluated for cardiovascular disease prevention should undergo 10-year atherosclerotic cardiovascular disease (ASCVD) risk estimation and have a clinician–patient risk discussion before starting on pharmacological therapy, such as antihypertensive therapy, a statin, or aspirin. In addition, assessing for other risk-enhancing factors can help guide decisions about preventive interventions in select individuals, as can coronary artery calcium scanning.”

In this study, we focus on coronary artery disease (CAD), a subcomponent of cardiovascular disease (CVD), as the prevalence of premature CAD is significantly higher than that of premature atherosclerotic stroke [9,10], and thus a focus for prevention.

**Hypertension**
USPSTF guidelines [11] state that: “The USPSTF recommends screening for hypertension in adults 18 years or older with office blood pressure measurement (OBPM).”

**Prostate cancer**
USPSTF guidelines [12] state that: “For men aged 55 to 69 years, the decision to undergo periodic prostate-specific antigen (PSA)-based screening for prostate cancer should be an individual one. Before deciding whether to be screened, men aged 55 to 69 years should have an opportunity to discuss the potential benefits and harms of screening with their clinician and to incorporate their values and preferences in the decision. Screening offers a small potential benefit of reducing the chance of death from prostate cancer in some men. However, many men will experience potential harms of screening, including false-positive results that require additional testing and possible prostate biopsy; overdiagnosis and overtreatment; and treatment complications, such as incontinence and erectile dysfunction. Harms are greater for men 70 years and older. In determining whether this service is appropriate in individual cases, patients and clinicians should consider the balance of benefits and harms on the basis of family history, race/ethnicity, comorbid medical conditions, patient values about the benefits and harms of screening and treatment-specific outcomes, and other health needs. Clinicians should not screen men who do not express a preference for screening and should not routinely screen men 70 years and older.”

The USPSTF guidelines recommend men aged 55 to 69 years should have an opportunity to discuss the potential benefits and harms of screening with their clinician, and that patients and clinicians should consider the balance of benefits and harms on the basis of family history, race/ethnicity, comorbid medical conditions, patient values about the benefits and harms of screening and treatment-specific outcomes, and other health needs. These guidelines are based on mixed results from prior randomised controlled trials on screening efficacy. These trials included all men, regardless of prior risk. The USPSTF guidelines state that family history of prostate cancer is one of the most important risk factors for prostate cancer and should be included in risk discussions and decisions. Given all these above factors, we assumed that a high PRS for prostate cancer would constitute a risk factor, and patients would be open to screening and more at risk of premature disease.

**Type 2 diabetes**
USPSTF guidelines [13] state that: “The USPSTF recommends screening for prediabetes and type 2 diabetes in adults aged 35 to 70 years who are overweight or obesity. Clinicians should offer or refer patients with prediabetes to effective preventive interventions.”

Being overweight or obese is a risk factor for type 2 diabetes. The risk inferred from obesity is similar to that from high PRS [14–16]. We assume that selective screening for type 2 diabetes would also be motivated for men at high risk of type 2 diabetes due to their PRS.

#### Definition of diseases for PRS effect size estimation

We used the following disease definitions within UK Biobank to define cases for the purposes of estimating PRS effect size (and also for the sensitivity analyses that employed empirical case events in UK Biobank rather than national incidence data).

| Disease | ICD-10 codes | Other requirements for UKB, in addition to Hospital Episode Statistic (HES) report with specified ICD-10 code |
|---|---|---|
| Abdominal aortic aneurysm | I713, I714 | Hospital operation codes L184, L185, L186, L194, L195, L196, L271, L275, L276, L281, L285, L286 |
| Breast cancer | C50 | Invasive breast cancer as reported by Cancer Registry or HES (ICD10 code C50, ICD9 code 174). Self-reported cancers (UKB code 1002 in field 20001) without a health record report of invasive breast cancer were excluded from analyses. Males set to 'missing' for GWAS and PRS evaluation. |
| Colorectal cancer | C18, C19, C20 | Small intestine/small bowel cancer, large bowel cancer/colorectal cancer, colon cancer/sigmoid cancer or rectal cancer as reported by Cancer Registry or HES (ICD10 codes C18, C19, C20; ICD9 codes 153, 154). Self-reported cancers (UKB codes 1019, 1020, 1022, 1023 in field 20001) without a health record of bowel cancer were excluded from analyses. |
| Coronary artery disease | I21, I22, I23, I24.1, I25.2 | ICD9: 410-412 and 42979, OPCS-4 codes (K40.1–40.4, K41.1–41.4, K45.1–45.5,K49.1–49.2, K49.8–49.9, K50.2, K75.1–75.4, K75.8–75.9), self-reported heart attack (UKB codes 1075 in field 20002; code 1 in field 6150) and self-reported coronary angioplasty (ptca) or coronary artery bypass graft (UKB codes 1070 and 1095 in field 20004). ICD10 codes were obtained both from hospital records and death records. |
| Hypertension | I10, I15 | Self-reported hypertension or essential hypertension (UKB codes 1065, 1072 in field 20002). |
| Prostate cancer | C61 | Prostate cancer as reported by Cancer Registry or HES (ICD10 code C61; ICD9 code 185). Self-reported prostate cancer (UKB code 1044 in field 20001) without a health record of prostate cancer were excluded from analyses. Male only. |

| | | |
|---|---|---|
| Type 2 diabetes | E11 | For prevalent cases, as a first step a decision tree approach previously described in Eastwood et al 2016 [PMID:27631769] was used to group individuals into different categories of likelihod of T1D or T2D. For individuals with GP data, we defined as likely T2D if they only had diagnosis codes mapping unambiguously to E11 (ICD10) or if any code mapping to E10 (ICD10) was present more than one year after a code mapping to E11 (ICD10). Individuals were defined as prevalent T2D if: a) they were likely T2D according to GP data; b) they were probable T2D according to decision tree and did not have GP data; or c) if there was a hospital record for E11 (ICD10) preceding date of assessment and they were unlikely T1D (by decision tree or GP data). Incident cases were defined by either unambiguous GP/HES code mapping to E11 (ICD10) only, or if any code mapping to E10 (ICD10) in GP/HES was present more than one year after a code mapping to E11 (ICD10). Individuals we could not confidently define as T2D or T1D were excluded from analysis. |

#### Estimation of disease characteristics and screening effects

##### Abdominal aortic aneurysm

Guirguis-Blake et al [17] present an evidence report and systematic review on screening for abdominal aortic aneurysm (AAA) in men, for the United States Preventive Services Taskforce. From their Table 1, a pooled odds ratio estimate of 0.65 (95% CI 0.57-0.74) is given for the relative AAA-based mortality rates in men in the screened versus unscreened arms of four randomised controlled trials (Chichester, Viborg, MASS, and Western Australia). Note this is an estimate of the odds ratio rather than the relative risk; since death rates are low (see their Figure 3), these estimates may be considered equivalent. All four randomised controlled trials were conducted on men regardless of their smoking status or history. The eligible age range varied from 64 to 83.

From Table 1 and Figure 3 of Guirguis-Blake et al, the number of AAA cases detected in the screened arms of the 4 studies is 228+247+1660+1386 = 3521, while the total sample size in the screened arms of the 4 studies is 2995+6333+33883+19249 = 62460, which generates an AAA prevalence under screening of 3521/62460 = 0.056. From Figure 3 of Guirguis-Blake et al, the number of AAA-related deaths in the unscreened arms of the 4 studies is 54+381+55+98 = 588, while the total sample size in the screened arms of the 4 studies is 3045+33887+6303+19231 = 62466, which generates an unconditional AAA mortality risk under no screening of 588/62466 = 0.009. Assuming that the AAA prevalence under screening is the same as the AAA prevalence under no screening, the conditional probability of AAA-related death, given one is a AAA case, is (588/62466)/(3521/62460) = 0.167, with 95% CI based on the Wald interval of 0.153 - 0.182. Note that this is a conservative underestimate, since the rate of AAA case detection in the unscreened arms will be lower than the rate of detection in the screened arms.

##### Female breast cancer

Henderson et al [18] present an evidence report and systematic review on screening for breast cancer in women, for the United States Preventive Services Taskforce, but this review does not directly report on the relative efficacy of screening versus no screening. However, Duffy et al [19] report a randomised controlled trial in the UK (UK Age trial), comparing mortality rates in women offered mammogram screening starting at age 40 to women offered mammogram screening starting at age 50 (the standard of care in the UK). The relative risk of mortality in the intervention versus control arms, arising from cancers detected in the ~10 year period between the start of the trial at age 40 and the commencement of standard screening at age 50, is 0.88 (95% CI 0.74–1.03) (Table 1 of Duffy et al).

Nelson et al [20] present an evidence report and systematic review on medication use for the risk reduction of primary breast cancer in women, for the United States Preventive Services Taskforce. Pooled results from 4 randomised controlled trials (n = 28,421) on the prophylactic use of tamoxifen indicate an invasive breast cancer risk ratio of 0.69 (95% CI 0.59–0.84).

From Table 2 of Duffy et al, in the control group (n = 106,953), 1628 breast cancer cases were detected via standard of care in the ~10 year period between the start of the trial at age 40 and the commencement of standard screening at age 50. Disease-specific mortality occurred in 474 of these cases during median 22.8 years follow-up (Table 1 of Duffy et al), leading to a conditional probability of breast cancer-related death, given one is a case, of 474/1628 = 0.291, with 95% CI based on Clopper-Pearson confidence intervals of 0.269–0.314.

**Colorectal cancer**

Lin et al [21] present an evidence report and systematic review on screening for colorectal cancer, for the United States Preventive Services Taskforce. Pooled results from 4 randomised controlled trials (n = 458,002) on flexible sigmoidoscopy compared with no screening indicate a mortality rate ratio of 0.74 (95% CI 0.68–0.80), over 11 to 17 years of follow-up.

Nishihara et al [22] use data from two prospective cohort studies: the Nurses' Health Study, which included 121,700 U.S. female nurses, 30 to 55 years of age at enrollment in 1976; and the Health Professionals Follow-up Study, which included 51,529 U.S. male health professionals, 40 to 75 years of age at enrollment in 1986. A "no lower endoscopy" group (n = 14,287 men, n = 31,423 women, their Table 1) was defined retrospectively from questionnaire data, to define an unscreened group. In this group, 1,164 cases of colorectal cancer were observed (their Table 2), and 349 colorectal cancer-related deaths (their Table 4), leading to a conditional probability of colorectal cancer-related death, given one is a case, of 349/1164 = 0.300, with 95% CI based on Clopper-Pearson confidence intervals of 0.274–0.327.

**Coronary artery disease**

Chou et al [23] present an evidence report and systematic review on primary prevention for cardiovascular disease, for the United States Preventive Services Taskforce. These authors do not report risk ratios for our definition of coronary artery disease (myocardial infarction and/or fatal coronary heart disease). We therefore used a previous meta-analysis of 14 randomised trials of statins for primary prevention by Baigent et al [24]. Baigent et al report a relative risk for myocardial infarction or coronary death of 0.77 (95% CI 0.74–0.80).

From eFigure 5 of Chou et al, the number of fatal myocardial infarction events reported in the untreated arms of 6 randomised controlled trials of statins for primary prevention (ALLHAT-LLT (1ry prevention) n=4405, CAIUS n=154, CARDS n=1410, JUPITER n=8901, KAPS n=212, and MEGA n=3966) is 96. From eFigure 2 of Chou et al, the number of fatal or non-fatal myocardial infarction event reported in the untreated arms of the same 6 trials is 216+2+61+68+8+33 = 388, leading to a conditional probability of coronary artery disease-related death, given one is a case, of 96/388 = 0.247, with 95% CI based on Clopper-Pearson confidence intervals of (0.205–0.293). Note that this is a conservative underestimate, as fatal coronary heart disease events not related to myocardial infarction are omitted.

**Hypertension**
Guirguis-Blake et al [25] present an evidence report and systematic review on screening for hypertension, for the United States Preventive Services Taskforce. Their review cites one randomised controlled trial for the effect of screening for hypertension on relevant outcomes (CHAP trial, Kaczorowski et al [26]). CHAP is a study of 39 mid-sized communities in Ontario, Canada, stratified by location and population size. Communities were randomised to receive Cardiovascular Health Awareness Program (CHAP) (n=20) or no intervention (n=19). In CHAP communities, residents aged 65 or over were invited to attend volunteer run cardiovascular risk assessment and education sessions held in community based pharmacies over a 10 week period; automated blood pressure readings and self reported risk factor data were collected and shared with participants and their family physicians and pharmacists. Kaczorowski et al report a relative risk for in-hospital cardiovascular mortality of 0.86 (95% CI 0.73–1.01).

From Table 3 of Kaczorowski et al, the probability of in-hospital death from cardiovascular disease in the control arm of the trial is 0.00466. Kaczorowski et al do not report the observed prevalence of hypertension in either arms of the study. However, data from NHANES [27] indicates a prevalence of hypertension in the USA in adults aged 60+ of 74.5%. Assuming that the prevalence of hypertension in the USA and Canada is similar, and that the prevalence in adults aged 65+ is similar to 60+, the conditional probability of hypertension-related death, given one is a case, can be estimated as 0.00466/0.745 = 0.0063, with 95% CI based on Clopper-Pearson confidence intervals of 0.0056–0.0069.

**Prostate cancer**
Fenton et al [28] present an evidence report and systematic review on prostate-specific antigen (PSA) screening for prostate cancer, for the United States Preventive Services Taskforce. Fenton et al note that, of three randomised controlled trials on the efficacy of PSA screening, one (ERSPC) provides a significant relative risk for prostate-specific mortality (0.79), while the other two (PLCO and CAP) do not. However, Tsodikov et al [29] present an analysis reconciling the results of ERSPC and PLCO, while a more recent report by Martin et al [30], over a 15 year follow-up period, finds a significant relative risk for prostate-specific mortality in CAP (0.92, 95% CI, 0.85-0.99). From the meta-analysis of ERSPC and PLCO by Tsodikov et al, the hazard ratio

for the effect of PSA screening for prostate cancer on prostate cancer-related mortality is 0.84, with 95% CI of 0.73–0.96. Since death rates are low, the hazard ratio and relative risk may be considered equivalent.

Martin et al [31] report the primary analysis of the CAP randomised controlled trial in the UK, evaluating the effect of a single PSA screening intervention and standardized diagnostic pathway on prostate cancer-specific mortality. The trial was conducted on 419 582 men aged 50 to 69 years, at 573 primary care practices across the United Kingdom. They report that the number diagnosed with prostate cancer in the control group was 7853, and that, after a median follow-up of 10 years, 647 had died of prostate cancer in the control group, leading to a conditional probability of prostate cancer-related death, given one is a case, of 647/7853 = 0.082, with 95% CI based on Clopper-Pearson confidence intervals of 0.076–0.089.

**Type 2 diabetes**

Jonas et al [32] present an evidence report and systematic review on screening for type 2 diabetes (T2D) for the United States Preventive Services Taskforce. From their Table 2, from a pooled analysis of 15 randomised controlled trials on the benefits of pharmacologic treatment for individuals with prediabetes, the relative risk of progression to diabetes under metformin treatment is 0.73 (95% CI 0.64–0.83). We assume that regular screening within the at-risk PRS group would enable effective identification of prediabetes before progression to diabetes, and thus an opportunity in all cases for prophylactic intervention with metformin.

From Figure 3 of Jonas et al, the number of T2D-related deaths reported in the untreated arms of 6 randomised controlled trials for various interventions on individuals recently diagnosed with diabetes is 62+129+55+122+297+120 = 785, while the sample sizes in those untreated arms was 328+1009+356+268+841+291+62+129+55+122+297+120 = 3878, leading to a conditional probability of T2D-related death given being a case of 785/3878 = 0.202, with 95% CI based on Clopper-Pearson confidence intervals of 0.190–0.215.

#### Definition of risk groups

Assuming a log-linear relationship between PRS and OR, and assuming a standard normal distribution for the PRS, and thus a log-normal distribution for the OR, the relationship between OR (relative to population mean risk) and PRS is given by the following equation. The $\beta^2/2$ correction is required for the arithmetic average of the population OR to be equal to one [33].

$$OR(x) = \exp(\beta . x - (\beta^2/2))$$

where x is the PRS value and β is the log of the OR per 1 SD of the PRS. For each disease, this formula was used to find the smallest PRS value such that the OR would be >2, or >3, and hence to assign individuals within the testing set with a PRS above the threshold to the relevant risk group. Note that the ‘high risk’ group includes all members of the ‘very high risk’ group.

##### Population disease incidence and mortality rates

US national disease incidence rates by age, sex and self-reported race/ethnicity were used both to estimate the risk advanced period, and to estimate the number of disease events expected during that period. Self-reported race/ethnicity groupings followed US Census Bureau derived designations of Non-Hispanic White, Non-Hispanic Black, Non-Hispanic Asian/Pacific Islander, Non-Hispanic American Indian/Alaska Native, and Hispanic. For breast cancer, colorectal cancer, and prostate cancer, incidence rates were obtained from the NIH Surveillance, Epidemiology, and End Results (SEER) program resource (2014-2018) [34]. SEER registries that report data are upheld to data standards as part of the North American Association of Central Cancer Registries (NAACCR). The completeness standard for malignant primary tumors is 98 percent or greater [35]. For coronary artery disease, incidence rates were obtained from heart disease statistics reported by the American Heart Association (2015 update) [36]. For hypertension, incidence rates were obtained from the National Health and Nutrition Examination Survey (NHANES) [37]. Annually, NHANES examines a nationally representative sample of 5,000 persons of all ages. To produce reliable statistics, the survey oversamples non-Hispanic black and Hispanic persons, and persons aged 60 and over. Physical examinations are conducted in mobile examination centers that travel to 15 U.S. counties annually. Participants aged 18 years and older (n = 4,689) in the National Health and Nutrition Examination Survey (NHANES 2017–2018) had their BP measured following 2 protocols: the legacy auscultation protocol (AP) and oscillometric protocol (OP) [38]. For type 2 diabetes, incidence rates were obtained from CDC National Health Interview Survey questionnaires (2015-2019) [39]. For abdominal aortic aneurysm, we were unable to identify suitable US population incidence rates for AAA: we therefore used male incidence rates derived from the UK Biobank. US disease specific mortality and all-cause mortality rates were obtained from CDC WONDER [40].

UK national disease incidence rates were obtained for selected diseases (sensitivity analysis) as follows. For breast cancer, colorectal cancer, and prostate cancer, incidence rates were obtained from the Office for National Statistics (ONS) cancer registration statistics, based on data collected in England in 2017 [41], and from Cancer Research UK [42]. For type 2 diabetes, incidence rates were obtained from Hippisley-Cox & Coupland [43]. UK disease specific mortality and all-cause mortality rates were obtained from the UK ONS cause of death survey [44].

3. Kelemen M, Danesh J, Di Angelantonio E, Inouye M, O'Sullivan J, Pennells L, et al.

Evaluating the cost-effectiveness of polygenic risk score-stratified screening for abdominal aortic aneurysm. Nat Commun. 2024;15: 8063. doi:10.1038/s41467-024-52452-w

15. Shoaib M, Ye Q, IglayReger H, Tan MH, Boehnke M, Burant CF, et al. Evaluation of polygenic risk scores to differentiate between type 1 and type 2 diabetes. Genet Epidemiol.

2023;47: 303–313. doi:10.1002/gepi.22521

doi:10.1136/bmj.d442

34. SEER*Explorer: An interactive website for SEER cancer statistics. Available: https://seer.cancer.gov/statistics-network/explorer/.

35. Surveillance, Epidemiology, and End Results Program (SEER) - Healthy People 2030 | odphp.health.gov. [cited 22 Jun 2025]. Available: https://odphp.health.gov/healthypeople/objectives-and-data/data-sources-and-methods/data-sources/surveillance-epidemiology-and-end-results-program-seer

36. Mozaffarian D, Benjamin EJ, Go AS, Arnett DK, Blaha MJ, Cushman M, et al. Heart disease and stroke statistics--2015 update: a report from the American Heart Association. Circulation. 2015;131: e29-322. doi:10.1161/CIR.0000000000000152

37. CDC National Center for Health Statistics. National Health and Nutrition Examination Survey (NHANES). Available: https://wwwn.cdc.gov/nchs/nhanes/Default.aspx

38. Ostchega Y, Hughes JP, Kit B, Chen T-C, Nwankwo T, Commodore-Mensah Y, et al. Differences in Hypertension and Stage II Hypertension by Demographic and Risk Factors, Obtained by Two Different Protocols in US Adults: National Health and Nutrition

Examination Survey, 2017–2018. Am J Hypertens. 2022;35: 619–626. doi:10.1093/ajh/hpac042

44. UK Office for National Statistics. Deaths registered in England and Wales – 21st century mortality. In: https://www.ons.gov.uk/peoplepopulationandcommunity/birthsdeathsandmarriages/deaths/datasets/the21stcenturymortalityfilesdeathsdataset.

### Supplementary Figures

#### Figure S1

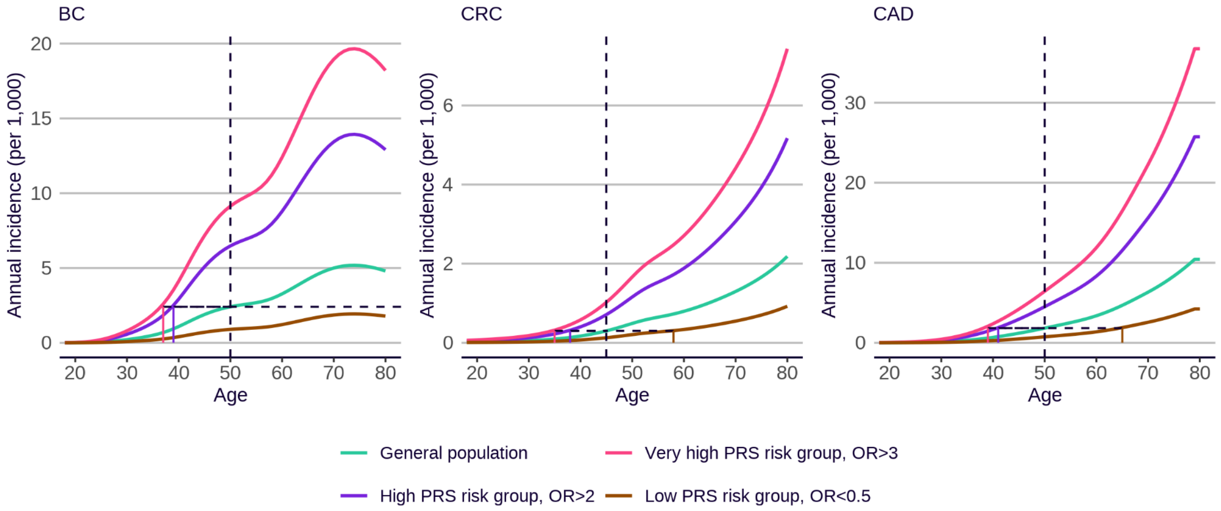


Figure S1. Sensitivity analyses using alternative conventional screening ages for breast cancer (BC) (age 50), colorectal cancer (CRC) (age 45), and coronary artery disease (CAD) (age 50). Panels show annual incidence for individuals at high (PRS OR>2), very high (PRS OR>3), and low risk (PRS OR<0.5) compared with the general population, using incidence rates for White self-identified race/ethnicity and PRS effect sizes for European genetic ancestry. For diseases that are not sex-specific, incidences are sex-averaged. Vertical dotted lines indicate the conventional screening start age. Horizontal dotted lines indicate the RAPs for different PRS-based risk groups. BC = female breast cancer; CRC = colorectal cancer; CAD = coronary artery disease.

#### Figure S2

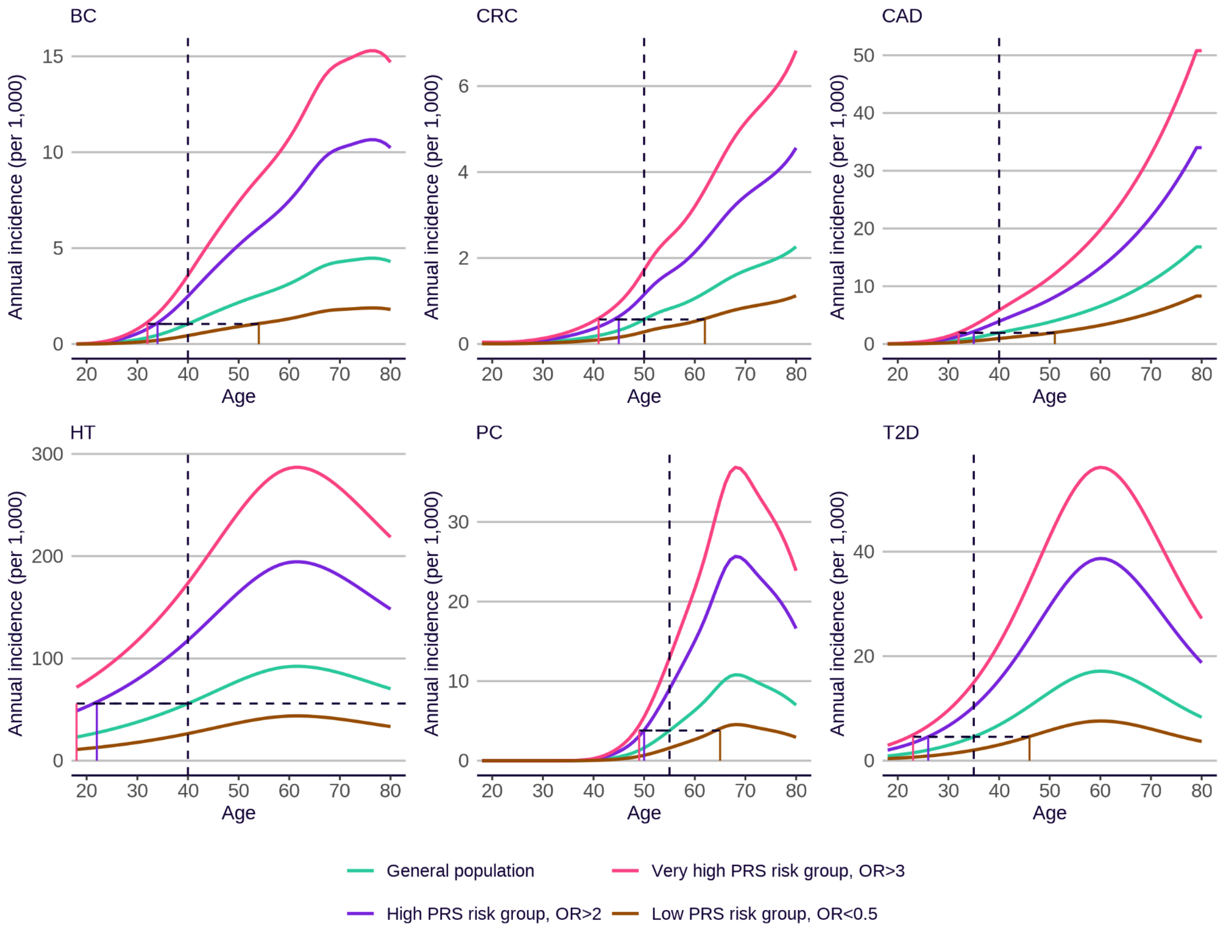


Figure S2. Sensitivity analyses using alternative incidence and PRS effect size data. Panels show annual incidence for individuals at high (PRS OR>2), very high (PRS OR>3), and reduced risk (PRS OR<0.5) compared with the general population, using incidence rates for Black self-identified race/ethnicity and PRS effect sizes for African genetic ancestry. For diseases that are not sex-specific, incidences are sex-averaged. Vertical dotted lines indicate the conventional screening start age. Horizontal dotted lines indicate the RAPs for different PRS-based risk groups. We were not able to estimate annual incidence for abdominal aortic aneurysm, due to the rates being too low in UK Biobank. BC = female breast cancer; CRC = colorectal cancer; CAD = coronary artery disease; HT = hypertension; PC = prostate cancer; T2D = type 2 diabetes.

#### Figure S3

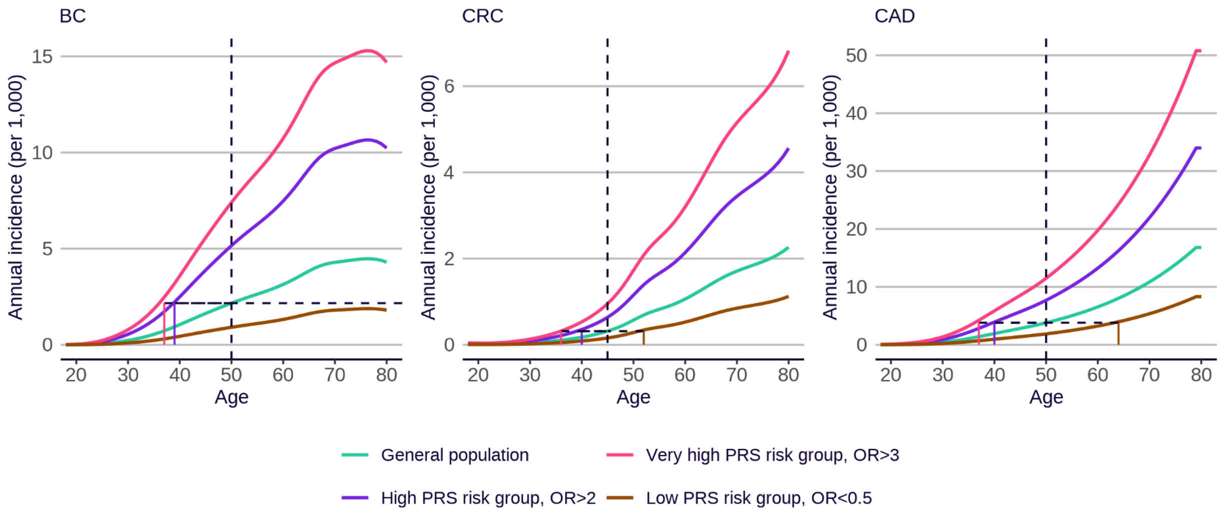


Figure S3. Sensitivity analyses using alternative incidence and PRS effect size data, and alternative conventional screening ages for breast cancer (BC) (age 50), colorectal cancer (CRC) (age 45), and coronary artery disease (CAD) (age 50). Panels show annual incidence for individuals at high (PRS OR>2), very high (PRS OR>3), and low risk (PRS OR<0.5) compared with the general population, using incidence rates for Black self-identified race/ethnicity and PRS effect sizes for African genetic ancestry. For diseases that are not sex-specific, incidences are sex-averaged. Vertical dotted lines indicate the conventional screening start age. Horizontal dotted lines indicate the RAPs for different PRS-based risk groups. BC = female breast cancer; CRC = colorectal cancer; CAD = coronary artery disease.

### Figure S4

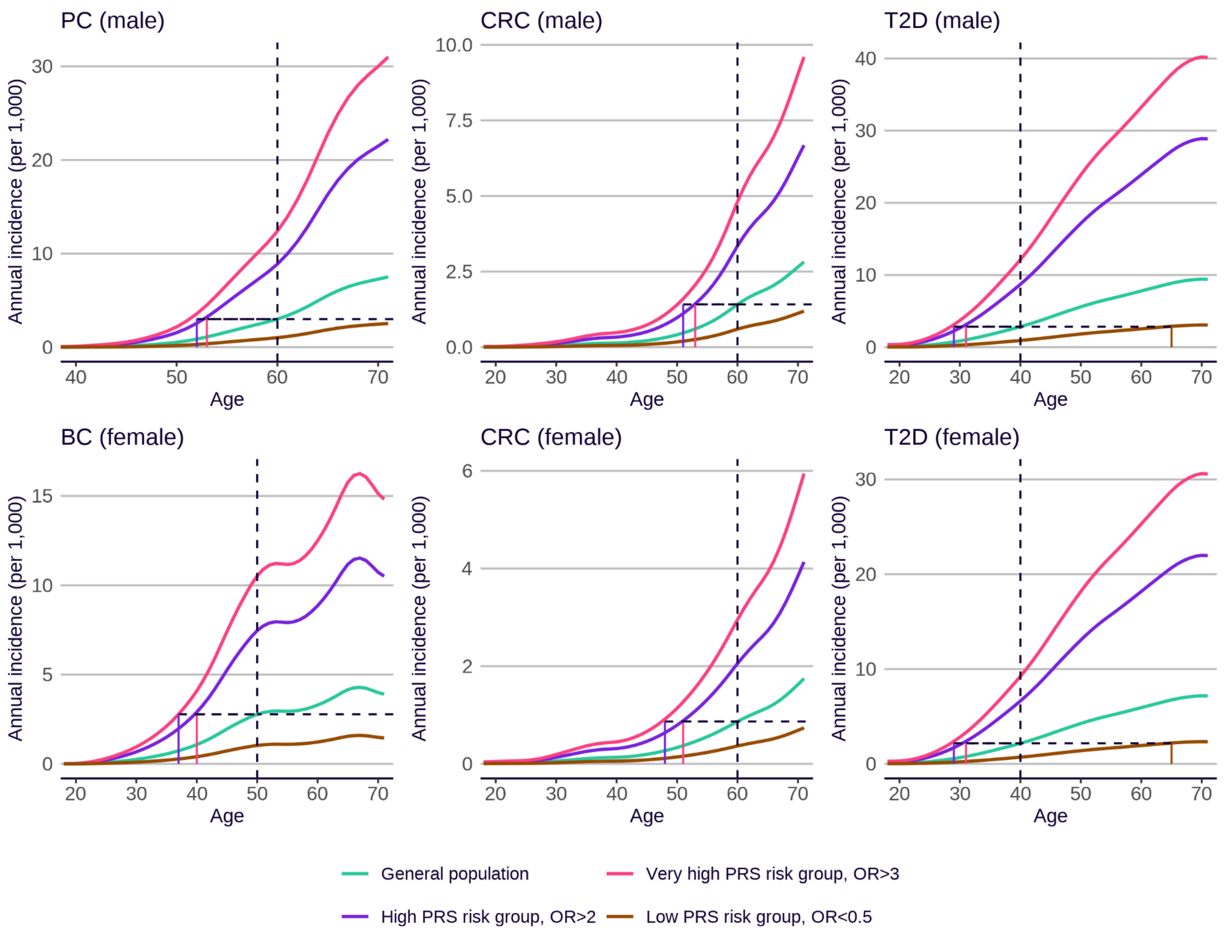


Figure S4. Sensitivity analyses using alternative incidence and PRS effect size data, and alternative conventional screening ages, based on UK data and guidelines. Panels show annual incidence with age for individuals at high (PRS OR>2), very high (PRS OR>3), and reduced risk (PRS OR<0.5), compared to the general population, using UK incidence rates for White self-identified race/ethnicity, and PRS effect sizes for European genetic ancestry. Vertical dotted lines indicate the conventional screening start age. Horizontal dotted lines indicate the RAPs for different PRS-based risk groups. BC = breast cancer (female); CRC = colorectal cancer; PC = prostate cancer; T2D = type 2 diabetes.
